# Toward adoption of health risk assessment in population-based and clinical scenarios

**DOI:** 10.1101/2023.08.02.23292593

**Authors:** Ruben Gonzalez-Colom, David Monterde, Roberta Papa, Mart Kull, Andres Anier, Francesco Balducci, Isaac Cano, Marc Coca, Marco De Marco, Giulia Franceschini, Saima Hinno, Marco Pompili, Emili Vela, Jordi Piera-Jiménez, Pol Pérez, Josep Roca, the JADECARE consortium

## Abstract

**Introduction:** Health risk assessment (HRA) strategies are cornerstone for health systems transformation toward value-based patient-centred care. However, steps for HRA adoption are undefined. This report analyses the process of transference of the Adjusted Morbidity Groups (AMG) algorithm from the Catalan Good Practice to the Marche region (IT) and to Viljandi Hospital (EE), within the JADECARE initiative (2020–2023).

**Description:** The implementation research approach involved a twelve-month pre-implementation period to assess feasibility and define the local action plans, followed by a sixteen-month implementation phase. During the two periods, a well-defined combination of experience-based co-design and quality improvement methodologies were applied.

**Discussion:** The evolution of the Catalan HRA strategy (2010–2023) illustrates its potential for health systems transformation, as well as its transferability. The main barriers and facilitators for HRA adoption were identified. The report proposes a set of key steps to facilitate site customized deployment of HRA contributing to define a roadmap to foster large-scale adoption across Europe.

**Conclusions:** Successful adoption of the AMG algorithm was achieved in the two sites confirming transferability. Marche identified the key requirements for a population-based HRA strategy, whereas Viljandi Hospital proved its potential for clinical use paving the way toward value-based healthcare strategies.

## BACKGROUND & OBJECTIVES

Health risk assessment (HRA) is a comprehensive approach that entails identifying, evaluating, and prioritising potential health risks and vulnerabilities for individuals and populations and identifying possible measures to reduce or mitigate their effects.

Deploying appropriate HRA strategies is a cornerstone for population risk stratification and constructing the corresponding population risk pyramid. It has become essential for informing health policy decisions, allocating resources, benchmarking, implementing preventive strategies and selecting appropriate healthcare services(1–3). Likewise, in the clinical arena, HRA is a building block for generating predictive models to support clinical decision-making(4,5). Indeed, population-based, and clinically-oriented HRA approaches are complementary elements needed to efficiently adopt integrated patient-centred care strategies. Deploying and adopting well-planned HRA strategies constitute an obligatory step toward the maturity of precision medicine(6).

Moreover, the momentum propitiated by the continuous progress in digital technologies for data capture and management, artificial intelligence and the advances in medical sciences are shaping novel and stimulating scenarios for health promotion and care and positioning predictive medicine at the forefront(7–9). Despite the promising potential of HRA, there is a noticeable gap between its benefits and its current application, attributed to several limitations, including the utilisation of suboptimal risk assessment tools, the insufficient engagement of health professionals, the application of ineffective or inexistent deployment strategies, and unresolved ethical and regulatory issues(10,11).

The Joint Action on implementation of digitally enabled integrated person-centred care (JADECARE)(12), an ongoing initiative launched to face the challenges of the transformation of health in the European Union, has included HRA as a strategic block to transfer from four original Good Practices (oGPs) to other twenty-one European regions participating as Next Adopters (NAs).

The central aims of JADECARE are to reinforce the capacity of health authorities for successfully addressing all the crucial aspects of health system transformation, in particular, the transition to digitally enabled, integrated, person-centred care, and to support the best practice transfer from the oGPs to the corresponding NAs. The current integrated case reports:

1. The evolution of the HRA strategy in one of the JADECARE oGPs (i.e., Catalonia, ES) from 2011-2020.
2. The description of the implementation process followed to transfer a population-based risk assessment tool from Catalonia to two NAs: Marche region (IT) and Estonia (EE).
3. The lessons learnt in the form of recommendations to foster the adoption of enhanced health risk assessment across the EU.

The report aims to identify key barriers and facilitators for effective adoption across Europe of population-based health risk assessment at the regional/country levels and formulate proposals facilitating the articulation between population-based and clinically-oriented HRA.

### DESCRIPTION OF THE CARE PRACTICE

#### HRA IN CATALONIA: 2011-2020

The 2011-2015 Catalan Health Plan(13) fostered key achievements in digital health transformation, the progressive implementation of person-centred integrated care services and the adoption of an initial population-based HRA strategy. This transitory HRA strategy was based on the commercial solution “Clinical Risk Groups” (CRG)(14) from vendor 3M™ and oriented toward modelling healthcare costs for resource allocation and benchmarking.

This initial HRA strategy laid the groundwork for a population-based risk stratification program with a case-finding approach focused on preventing adverse health events, managing high-risk chronic patients, and early detecting end-of-life patients(15,16). This HRA approach acknowledged the close relationship between frailty and multimorbidity while recognizing their distinct nature as independent risk factors. Additionally, it introduced specific scales for the evaluation of each factor individually. The clinical complexity level was assigned based on multimorbidity scoring, transitively using CRGs, and the clinical judgment of primary care physicians. Complex patients were classified into two groups: complex chronic patients (CCP), approximately 4% of the population, and advanced chronic patients (ACP), representing approximately 1% of the population with limited life expectancy. Specific community-based management plans, aiming at integrating health and social care, were defined for these two categories of patients(15). An overall description of the care model for people with frailty and multimorbidity in Catalonia, tested during the 2011-2015 Health Plan, has been recently reported in (17).

The need for refining the assessment of the multimorbidity burden triggered the creation of the Adjusted Morbidity Groups (AMG)(18,19), a new morbidity grouper that reflects patients’ disease burden in terms of the number and complexity of concomitant disorders through a disease-specific weighting deduced from statistical analysis based on mortality and the utilisation of healthcare resources. The AMG tool was jointly launched in 2015 by the Spanish Health Ministry and the Catalan public health commissioner (CatSalut). A significant achievement was the development of a dashboard to monitor the population’s multimorbidity burden and the use of healthcare known as Modules for Monitoring Quality Indicators (“Moduls pel Seguiment d’Indicadors de Qualitat”, MSIQ)(20), which generates and displays customised key performance indicators (KPIs) with aggregated data to inform health policy decisions, benchmarking, and governance. On the clinical side, the AMG scoring of the patient is currently displayed in the workstation of the primary care physicians and the shared clinical history(21) as a support tool in the clinical setting.

During the 2016-2020 Catalan Health Plan(22), the utilisation of AMG for HRA purposes was validated in different regions of Spain, covering a population of approximately 38 million citizens and showing good transferability in all cases(23). This period witnessed three significant advancements in Catalonia’s HRA strategy:

1. The execution of several studies testing the contribution of the AMG in different HRA settings, such as the identified risk factors during the SARS-CoV-2 pandemic(24,25), the refinement of tools for resources allocation(26–29), the analysis of the effect of the multimorbidity burden in patients with chronic obstructive pulmonary disease (COPD)(30), and assessing the use of AMG in complex clinical predictive models for short-term clinical outcomes predictions after hospital discharge(4,5).
2. The creation of the Catalan Health Information System Master Plan published in 2019(31) established the basis for the transformation to a new digital-health paradigm based on a knowledge-driven platform and adopting the Open-EHR standard as a reference.
3. The development and internal validation of the Queralt indices(32,33) to characterise the complexity of hospitalisation episodes, combining information on the principal discharge diagnosis, pre-existing comorbidities, in-hospital complications and all the procedures performed during hospitalisation.

Figure 1 depicts the information required from NAs to use the AMG, the outputs obtained, and the key uses of AMG.

**Figure 1.**
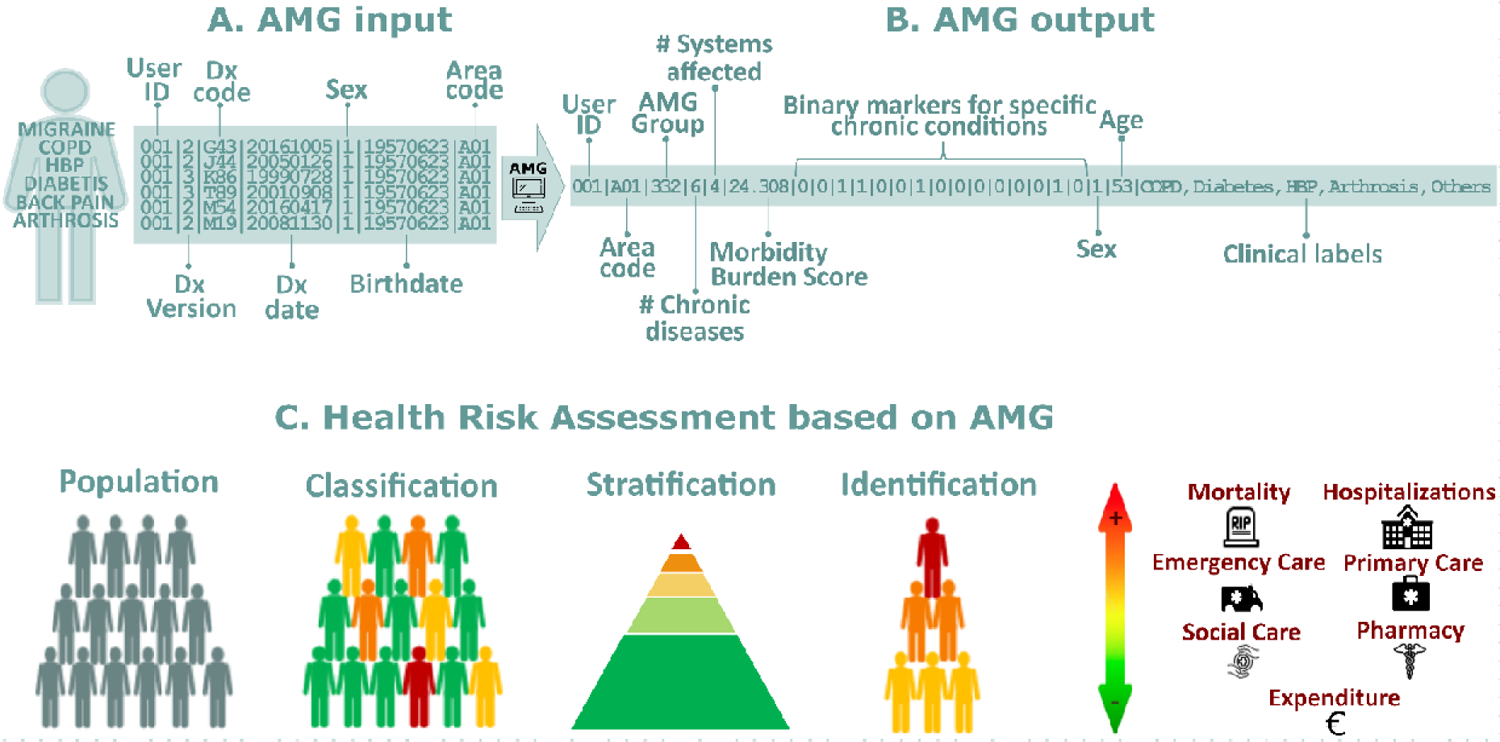
Panel A - AMG input: Required input variables to compute AMG; Panel B – AMG output: Output variables of the AMG algorithm. *Binary markers (presence/absence) of 15 chronic conditions (from left to right): diabetes mellitus, heart failure, chronic obstructive pulmonary disease, high blood pressure, depression, HIV/AIDS, chronic ischemic heart disease, stroke, chronic kidney disease, cirrhosis, osteoporosis, arthrosis, arthritis, dementia, chronic pain; Panel C– Health Risk Assessment based on AMG: The AMG scoring allows for three key actions: Classification: The population is categorised into specific groups based on their morbidity statuses, such as healthy, pregnancy and labour, acute disease, chronic disease in 1-4 systems, or active neoplasia, which are also divided into five degrees of severity. Stratification: Each individual can be assigned a complexity score that reflects the care needs that people may have based on their health problems. Identification: Individuals with specific major chronic health problems can be identified, which helps track people with more complex care needs.

### PRE-IMPLEMENTATION (October 2020 – September 2021)

Within JADECARE, the AMG was transferred to the Marche region and Estonia. The Marche region has a regionally based healthcare system, providing universal coverage to 1,480,839 citizens of which 25.4% are 65 years and older. Life expectancy is of 81 years for men and 85.2 for women. In comparison, Estonia has compulsory solidarity-based health insurance, financed by the health insurance budget through the Estonian Health Insurance Fund, covering 1,322,765 citizens, 29.0% aged 65 years and older. Life expectancy at birth is 72.8 years for men and 81.4 for women.

In Estonia, the initial implementation site for the AMG transfer was chosen was Viljandi County with approximately 30 general practitioners and one general hospital providing specialist care for around 50,000 inhabitants.

Table 1 describes the context and the trigger that motivated the adoption of the AMG in the Marche region and Estonia, alongside the local aims of each NA and the main challenges identified by the oGP leaders. Specific Local Action Plans were designed to fulfil the needs of each NA, as reported in detail in the Supplementary Material. The pre-implementation phase concluded once the implementation feasibility study was successful in each site.

**Table 1.** Summary of the pre-implementation process, including the context and trigger, the aims, the baseline situation, and the challenges faced in the Marche region and Estonia.

|  | Marche region | Estonia |
| --- | --- | --- |
| **Context and trigger** | 1) The high burden of non-communicable diseases (NCDs) and the need for more efficient management of affected patients.<br>2) The need for support decision-making in healthcare services and policies and analyse the utilisation of healthcare resources in alignment with national regulations defining population stratification as a prerequisite for healthcare planning (e.g. National Plan for Chronicity, 2016; National Decree on standards and organisation of community services, 2022). | 1) Digital infrastructure to support integrated care was piloted in Estonia with minimal impact and long-term traction, and there was not yet region-wide coverage. Likewise, social and healthcare service coordination is in an early phase.<br>2) The different and non-aligned payment schemes for hospital and ambulatory care impact incentivising the transformation from case-based care to a population health-oriented care model with social services integration to care.<br>3) Risk stratification and case-finding tools were needed to facilitate high-risk patient identification for regional care-management and service integration. |
| **Aims** | 1) To test and adopt the AMG population stratification algorithm, suitably adapted to the regional context and the available health data.<br>2) To display a regional dashboard for health policy purposes, benchmarking, and decision-making processes. | 1) To develop an integrated clinical program to prevent hospitalisations and target elders with concomitant chronic diseases and social health determinants.<br>2) To adopt the AMG for service selection.<br>3) To leverage the acquired expertise and progress towards innovative value-based reimbursement models, aiming to establish Viljandi Hospital as an accountable care organization.<br>4) To escalate the adoption of AMG for population stratification at country-level. |
| **Baseline situation** | The Regional Healthcare Administrative Databases (HADs), used at regional and national levels to monitor healthcare system expenditures and performance, gather information on healthcare services provided to citizens (e.g., hospitalisations, emergency-urgency, homecare, exemption codes, etc.). Common standard models and coding systems, such as DRGs(34) and ICD-9-CM(35), are used across all regions. However, each HAD has its unique structure, unit level, content, and rules for data input. Data linkage across HADs is facilitated using a unique anonymised patient ID code. | The Estonian Health Insurance Fund claims database was used for model data input. In Estonian universal healthcare and single payer model this database entails almost all medical care claims in the country. The database follows a single standard with ICD-10-CM(36) coding, DRGs. Data linkage and access was granted though ethics committee approval and is not easily available as standard. Data was analysed in anonymised format using unique patient ID codes. All AMG analyses were performed by the regional medical authorities under the ethics committee approved application with support from the oGP. |
| **Challenges** | 1) To fulfil the initial feasibility test.<br>2) To overcome potential technical problems in dynamically assembling the dataset required to feed the AMG algorithm using heterogeneous data sources.<br>3) The site showed an explicit limitation in the use of HRA tools due to the GDPR-related legislation at the Italian level regarding the secondary use of health data. | 1) To fulfil the feasibility test.<br>2) To ensure firm commitments of the Estonian government (Ministry of Social Affairs), as well as getting traction and commitment by the Estonian Health Insurance Fund to the project.<br>3) To overcome potential technical problems in dynamically assembling the dataset required to feed computational modelling for health risk stratification. |

### Results of the feasibility analysis

To assess the transferability of the AMG tool, independent feasibility tests were conducted in each of the adopting regions. To perform the AMG feasibility analysis in the Marche region, fully anonymized information on disease diagnostics from 2015 to 2019 was extracted from three independent databases: the hospital admissions, the emergency department, and the exemption codes for chronic and rare diseases databases. Integrating all the medical information resulted in 5,939,199 diagnostic codes associated with 1,367,181 citizens. In Estonia, the AMG feasibility study was conducted with 25,930 diagnostic codes related to 4,765 citizens treated in Viljandi Hospital in 2018.

The feasibility analysis evaluated the comprehensiveness of the clinical information accessed and the discrimination capacity of the AMG algorithm to identify high-risk individuals allocated in seven morbidity groups: healthy, pregnancy, acute disease, 1 or 2-3 or ≥ 4 chronic diseases, and active neoplasia.

When evaluating the feasibility study, it is imperative to consider the databases’ dual approach and purpose. The Marche database is a population health database(37), built for informing health policies, benchmarking, and supporting decision-making processes. On the other hand, the Viljandi database follows a population medicine approach(37), assembling patient registries to screen candidates for a clinical program geared towards preventing hospitalisations for elderly patients with concomitant chronic conditions.

The feasibility tests were deemed successful if the following conditions were met:

1. The algorithm effectively discriminated the patients with different risk profiles within specific age and gender groups, especially in older individuals more susceptible to multimorbidity.
2. The algorithm distinguished between the seven AMG disease groups. It’s worth noting that the relative frequencies of these groups may differ depending on the database’s nature.
3. The results showed a positive correlation between the clinical complexity of the AMG disease groups and the disease burden evaluated using the AMG Morbidity Burden Index.

The feasibility analysis results are shown in Figure 2. In Panel A, the distribution of the five subgroups of complexity is depicted according to age and gender. As expected, the demographic characteristics of the population treated in Viljandi utilised in this feasibility study differ from the Viljandi county’s population. Overall, the AMG algorithm proficiently distinguished patients with varying risk profiles in specific age and gender categories. Panel B shows the distribution of the seven AMG morbidity groups’ complexity level, represented by the average Morbidity Burden Score. In the Marche region and Estonia, roughly 50% of the population suffers at least one chronic disease and 20% developed multimorbidity, while 5% has an active neoplasm. Notably, the analysis of the population treated in Viljandi Hospital revealed a higher fraction of citizens with acute morbid conditions due to the hospital nature of the database. When comparing the relative distribution of AMG disease groups between the adopting regions and Catalonia, notable disparities emerge in terms of the prevalence of patients with multimorbidity. These variations can be attributed to the availability of primary care registries, which is a condition exclusively fulfilled in the Catalan context. This underscores the critical importance of integrating health data from diverse levels. The complexity of the disease groups and the AMG Morbidity Burden Score showed a strong positive correlation, being slightly higher in the Marche region due to the exhaustivity of the diagnostic records at different healthcare levels and the increased length of the study period.

**Figure 2.**
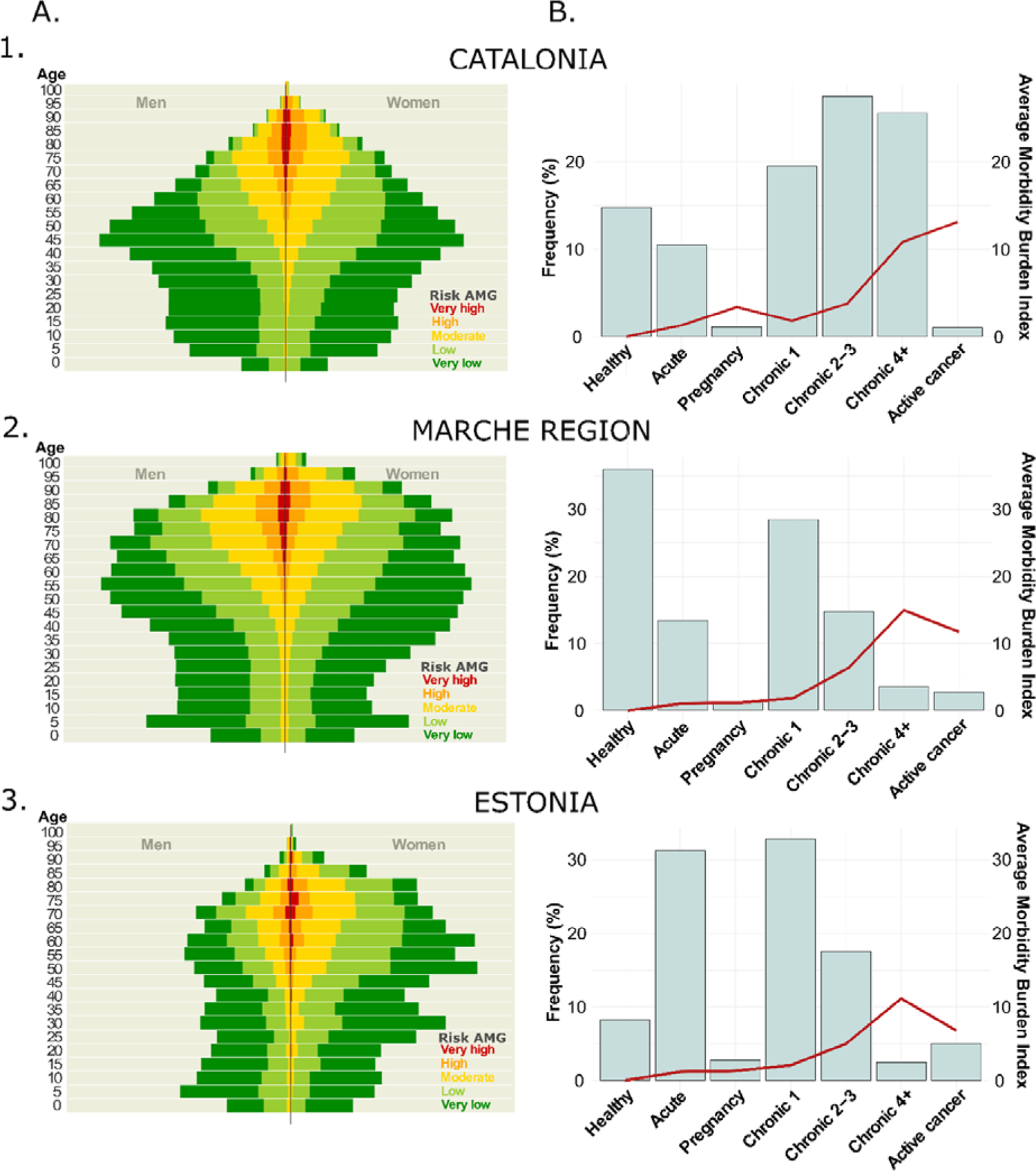
Results of the feasibility analysis: in Catalonia (1) and the adoption regions, Marche Region (2) and Estonia (3). Panel A - AMG risk distribution: itemised by age and gender; Panel B – AMG disease groups: distribution of the seven AMG morbidity groups (bars): healthy, pregnancy, acute disease, 1 or 2-3 or≥ 4 chronic diseases, and active neoplasia, and their average Morbidity Burden Score (line).

Based on the assessment made by the oGP specialists and the findings presented, it was determined that the databases generated by both adopters, the Marche region and Estonia, were mature and ready to expedite the implementation of the AMG.

### IMPLEMENTATION (October 2021-January 2023)

On October 2021, the Local Action Plans (LAPs) in Marche and Estonia were already available, and the corresponding NA Working Groups were prepared to undertake two Plan-Do-Study-Act (PDSA) cycles(38). The details of the PDSA cycles are reported in the Supplementary Material. A major midterm milestone of the implementation phase was the HRA Thematic Workshop, held in Viljandi Hospital on 14-15 June 2022. The workshop delimitated the bases for deploying population-based HRA strategies focused on AMG and the Queralt indices. A recording of the session can be found in the Supplementary Material.

#### Marche

Implementation achievements - 1) dynamic regional dataset preparation merging information from different existing data sources, 2) data cleaning and automatization of the generation of the regional dataset, 3) elaboration and analysis of the risk assessment pyramid at regional level demonstrating association between GMA scoring and local use of resources, 4) preparation of the logistics for local sustainability of the setting; and 5) design of the regional dashboard to facilitate regional health governance. The technicalities of the implementation and the assessment of the implementation process will be reported in a forthcoming paper.

Sustainability Action Plan - 1) to complete the integration of the tool into the regional IT infrastructure, adding further healthcare databases and defining supportive actions to improve the quality and completeness of healthcare data. 2) to implement the dashboard in computational, technical, and graphical terms, adding maps aimed to visualize healthcare services adjusted to social-health care planning regulations and integrating it in the regional IT infrastructure. 3) to promote the use of the HRA tools by regional and clinical managers and share our experience for the discussions on the secondary use of health data.

#### Estonia

Implementation achievements – 1) Generating a protocol, already approved by the local Ethics committee for a pragmatic randomized control trial (n= 1000), to test effectiveness and value generation of an integrated care intervention to prevent hospitalizations targeting community-based patients with high-risk of admissions and enhancing transitional care post-discharge to reduce early readmissions (PAIK 2022-2025). 2) AMG will be used as inclusion criteria and to modulate the characteristics of the intervention and the Queralt indices will be employed to characterize hospitalization episodes contributing to personalize transitional care after discharge.

Sustainability Action Plan - 1) to successfully execute the PAIK 2022-2025 project using AMG and Queralt indices as risk assessment tools.2) to generate sound proposals for innovative reimbursement modalities.3) to achieve country-wide scalability of risk prediction approaches based on AMG and Queralt.4) to initiate a debate on the scalability of integrated care services involving innovative reimbursement modalities in Estonia.

### PERSPECTIVES BEYOND IMPLEMENTATION

The section describes the status of the HRA in Catalonia and briefly reports the overarching analysis of the process of transference to the two NAs, leading to recommendations for the generalisation of the case practice at the European level.

#### Evolution of the Catalan oGP during JADECARE

The 2020 Catalan Health Information System Master Plan has sparked ongoing technological innovations that offer significant opportunities in predictive modelling aiming at supporting clinical decision-making for healthcare professionals and provide patients with decision support tools to empower self-management(39–41). Also, specific initiatives have been launched to enhance transitional care and reduce early readmissions after hospital discharge(4,5). Moreover, efforts are devoted to the practicalities of adoption of such predictive modelling tools in real-world settings, involving: 1) training and continuous evaluation of clinical decision support systems embedded into integrated care services, 2) pragmatic use of implementation science tools to foster engagement of health professionals, and 3) ethical and regulatory aspects including refinement of the regional PADRIS program(42) for secondary use of health data.

In Catalonia, research, and innovation in HRA are currently focused on two target areas. Firstly, to address unmet needs associated with enhanced predictive modelling considering four potential sources of variables: 1) clinical information, 2) registry data with a population health approach, 3) patients’ self-reported information (PROMs and PREMs) and self-monitoring data, and 4) biomedical research data, as defined in (1). A key learning from the different studies done using the AMG algorithm since 2015 is that multimorbidity has a central explanatory role in health risk assessment, more than classical variables like age, which prompts the inclusion of the AMG scoring as a fix covariate in multisource predictive modelling.

A second field of research interest is the potential evolutions of AMG incorporating information on patients’ disease trajectories(43,44). Although these novel computational developments are still far from being applicable in clinical settings, they present exciting prospects or precision medicine in patients with NCDs.

#### Overarching analysis

The case practice identifies the process of transference and adoption of the AMG algorithm and a site-tailored dashboard using aggregated data as the two key components necessary for establishing fundamental HRA functionalities.

While Marche adopted a population-health approach considering the use of healthcare resources from the entire geographical area, Viljandi Hospital in Estonia implemented a more limited regional population-medicine orientation with a single hospital as a local integrator and lead in a regional health improvement initiative. As depicted in Figure 2, the differences in terms of sources and composition of input data had significant impacts on the distribution of AMG scoring among Catalonia (Figure 2 - Panel 1, population-health data from all healthcare tiers), Marche (Figure 2 - Panel 2, population-health data with poor representation of primary care) and Viljandi Hospital (Figure 2 - Panel 3, population-medicine approach based on hospital information). Accordingly, population-health databases, as those used in Marche region, must comprise information on disease diagnostics from all individuals within the region, gathered from various tiers of the healthcare system (e.g. primary care, community, hospital, etc.). It is expected to find a significant fraction of the healthy population and cases with transitory health problems, both concentrated in the youngest fraction of the population. The remaining population is expected to suffer at least chronicity and many of them to develop multimorbidity; among them is expected to find a small fraction of complex chronic patients.

On the other hand, if the analysis is conducted following a population medicine approach, as done in Viljandi, the sample should be representative of the population treated at the hospital during the period assessed, both in terms of demographic parameters and the clinical profiles of the patients. In light of this, a highly biased population is expected to be older than the hospital’s catching area average. Also, a substantial increase in the frequency of patients experiencing mainly acute and chronic conditions is anticipated.

Consequently, the composition of the input datasets plays a crucial role in modulating both purposes and suitability of adopting HRA strategies. In the current case practice, Marche’s approach was adequate to cover health policy aspects, resources allocation, benchmarking, and governance at the regional level, even though limitations due to GDPR constraints at the Italian level should be acknowledged. In contrast, the HRA orientation adopted by Viljandi Hospital was beneficial in the design process of the PAIK2 protocol (2023–2025), aiming at generating evidence of the effectiveness of an integrated care intervention to prevent hospitalisations in high-risk patients and transitional care. It is of note that the process of developing the case practice in the two sites has generated knowledge and skills that will be needed to elaborate future site-customised comprehensive HRA strategies, as described in the case of Catalonia.

In summary, the essential maturity requirements for sustainable HRA strategies are: 1) to achieve solid political commitment at the regional/country level fostering necessary interactions, top-down and bottom-up, among key stakeholders; 2) to have an essential digital maturity at the site level to satisfactorily pass the feasibility analysis; 3) to overcome potential limitations due to local application of GDPR; 4) to use highly applicable implementation science tools fostering engagement of all stakeholders, health policy managers and health professionals, during the process elaboration and deployment of the local HRA strategy; and 5) to develop/adopt a regulatory framework for secondary use of health data, needed for business intelligence and the elaboration of computational modelling for clinical applications. Table 2 depicts the steps identified during the transference process to define a roadmap for adopting site-customised HRA strategies.

**Table 2.**
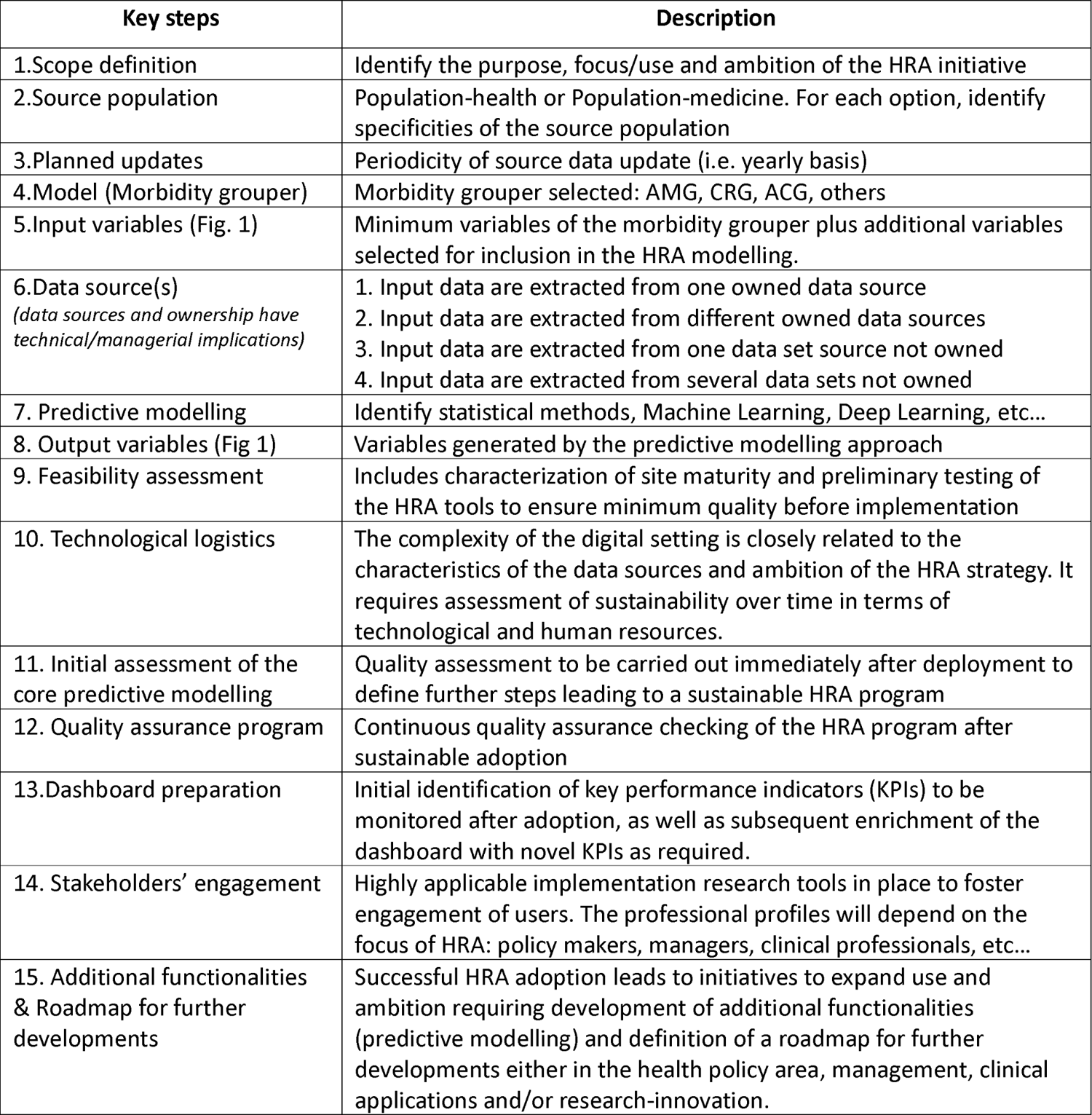
Checklist of key steps for site adoption of Health Risk Assessment (HRA)

| Key steps | Description |
| --- | --- |
| 1.Scope definition | Identify the purpose, focus/use and ambition of the HRA initiative |
| 2.Source population | Population-health or Population-medicine. For each option, identify specificities of the source population |
| 3.Planned updates | Periodicity of source data update (i.e. yearly basis) |
| 4.Model (Morbidity grouper) | Morbidity grouper selected: AMG, CRG, ACG, others |
| 5.Input variables (Fig. 1) | Minimum variables of the morbidity grouper plus additional variables selected for inclusion in the HRA modelling. |
| 6.Data source(s)<br>*(data sources and ownership have technical/managerial implications)* | 1. Input data are extracted from one owned data source<br>2. Input data are extracted from different owned data sources<br>3. Input data are extracted from one data set source not owned<br>4. Input data are extracted from several data sets not owned |
| 7. Predictive modelling | Identify statistical methods, Machine Learning, Deep Learning, etc... |
| 8. Output variables (Fig 1) | Variables generated by the predictive modelling approach |
| 9. Feasibility assessment | Includes characterization of site maturity and preliminary testing of the HRA tools to ensure minimum quality before implementation |
| 10. Technological logistics | The complexity of the digital setting is closely related to the characteristics of the data sources and ambition of the HRA strategy. It requires assessment of sustainability over time in terms of technological and human resources. |
| 11. Initial assessment of the core predictive modelling | Quality assessment to be carried out immediately after deployment to define further steps leading to a sustainable HRA program |
| 12. Quality assurance program | Continuous quality assurance checking of the HRA program after sustainable adoption |
| 13.Dashboard preparation | Initial identification of key performance indicators (KPIs) to be monitored after adoption, as well as subsequent enrichment of the dashboard with novel KPIs as required. |
| 14. Stakeholders' engagement | Highly applicable implementation research tools in place to foster engagement of users. The professional profiles will depend on the focus of HRA: policy makers, managers, clinical professionals, etc... |
| 15. Additional functionalities & Roadmap for further developments | Successful HRA adoption leads to initiatives to expand use and ambition requiring development of additional functionalities (predictive modelling) and definition of a roadmap for further developments either in the health policy area, management, clinical applications and/or research-innovation. |

Moreover, using the HRA tools described in the case practice as open-source software and articulating public-private collaborations supporting productive interactions and networking among sites are essential to speed up transferability and large-scale adoption of HRA strategies at the European level.

## DISCUSSION

The current report is filling an existing gap in information on building up an HRA strategy at the regional/country level, as described for Catalonia, and its potential for transferability to other sites. Moreover, the Marche and Viljandi Hospital process generated a helpful checklist for generalising such transference to other sites across Europe.

The case practice has also recognised the convenience of further collaboration among regions beyond JADECARE to keep progressing toward mature HRA strategies contributing to paving the way for precision medicine. Moreover, the use of the current and future available algorithms (i.e. AMG, Queralt, etc..), and dashboards, as open-source software, as well as the provision of consultancy services supporting future next adopters, were identified as core elements to foster the adoption of efficient HRA strategies across Europe. To this end, the elaboration of a survey to be administered to all JADECARE’s sites is strongly encouraged. The main aim is characterising the status and needs of all NAs regarding HRA. The information from the survey gathered before the project end can contribute to refining a large-scale implementation protocol to be customised at the site level beyond the project lifetime.

### LESSONS LEARNED

The analysis of the HRA strategies in Catalonia (2010–2023), as well as the process of transference and adoption of the AMG in Marche (IT) and Viljandi Hospital (EE) within JADECARE, 2020-2023, generated the following key learnings aiming at fostering large-scale adoption of HRA across Europe:

1. Adopting comprehensive HRA strategies, including multimorbidity weight (AMG scoring) as a central component, constitutes a key element fostering health systems transformation toward value-based healthcare with a patient-centred approach.
2. Population-based and clinically-oriented HRA must be considered complementary and highly synergistic. However, ethical and regulatory aspects for secondary use of health data must be appropriately assessed and locally implemented.
3. Transferability of AMG across sites with diverse source data models is feasible, provided that the key input variables are available, the source population is well-characterized, and adequate data management with quality assurance over time is in place.
4. The current analysis identified the relevant steps to be considered for a generic protocol aiming at the implementation of HRA at a regional level.
5. Short-term elaboration of a map describing both maturity levels and needs for HRA adoption in the twenty-one JADECARE sites could provide the necessary information to define a roadmap leading to large-scale implementation of HRA across Europe.

## CONCLUSIONS

The current study describes the evolution and the potential of the HRA strategy adopted in Catalonia since 2010. It also illustrates the transferability of the AMG algorithm in two different scenarios. Moreover, the case practice identified relevant barriers/facilitators modulating the adoption of HRA at regional level and elaborated a set of key steps to be considered for site deployment of HRA. Last, but not least, the report proposes initial steps to define a roadmap aiming at fostering large-scale implementation of HRA across Europe.

## ETHICAL APPROVAL

This research exclusively employed fully anonymized retrospective health records sourced from administrative databases. Consequently, in compliance with prevailing legislation in the regions where the analyses were conducted, neither informed consent nor ethical committee approval was required. All the analytical procedures were undertaken under the auspices of the JADECARE project.

## Data Availability

Due to the nature of the research and to ethical aspects, supporting data is not available.

## ACKNOWLEDGEMENTS

This research was funded by JADECARE project-HP-JA-2019 - Grant Agreement n° 951442 a European Union’s Health Program 2014-2020.

## ACRONYMS

ACP: Advanced Chronic Patients

AMG: Adjusted Morbidity Groups

CCP: Complex Chronic Patients

COPD: Chronic Obstructive Pulmonary Disease

CRG: Clinical Risk Groups

GDPR: General Data Protection Regulation

HAD: Healthcare Administrative Database

HRA: Health Risk Assessment

KPI: Key Performance Indicator

LAP: Local Action Plan

MSIQ: Modules for Monitoring Quality Indicators (“Moduls pel Seguiment d’Indicadors de Qualitat”)

NA: Next Adopter

NCD: Non-Communicable Disease

oGP: Original Good Practice

PDSA: Plan-Do-Study-Act

PREM: Patient Reported Experience Measure

PROM: Patient Reported Outcome Measure

## ON-LINE SUPPL MATERIAL

1. Local Action Plans
2. Plan-Do-Study-Act reports
3. The lecture by David Monterde in Viljandi – 14 June 2022 (video)

## REFERENCES

1. Dueñas-Espin I, Vela E, Pauws S, Bescos C, Cano I, Cleries M, et al. Proposals for enhanced health risk assessment and stratification in an integrated care scenario. BMJ Open. 2016 Apr 1;6(4):e010301.

2. Cano I, Dueñas-Espín I, Hernandez C, De Batlle J, Benavent J, Contel JC, et al. Protocol for regional implementation of community-based collaborative management of complex chronic patients. npj Prim Care Respir Med. 2017;27(1):1–6.

3. Brilleman SL, Gravelle H, Hollinghurst S, Purdy S, Salisbury C, Windmeijer F. Keep it simple? Predicting primary health care costs with clinical morbidity measures. J Health Econ. 2014;35:109–22.

4. Calvo M, González R, Seijas N, Vela E, Hernández C, Batiste G, et al. Health outcomes from home hospitalization: Multisource predictive modeling. J Med Internet Res. 2020 Oct 1;22(10):e21367.

5. González-Colom R, Herranz C, Vela E, Monterde D, Contel JC, Sisó-Almirall A, et al. Computational modelling for prevention of unplanned hospital admissions in multimorbid patients. J Med Internet Res. 2023;

6. Smith MA, Adelaine S, Bednarz L, Patterson BW, Pothof J, Liao F. Predictive Solutions in Learning Health Systems: The Critical Need to Systematize Implementation of Prediction to Action to Intervention. NEJM Catal Innov Care Deliv. 2021 Apr 21;2(5).

7. Bjerring JC, Busch J. Artificial Intelligence and Patient-Centered Decision-Making. Philos Technol. 2021;34:349–71.

8. He J, Baxter SL, Xu J, Xu J, Zhou X, Zhang K. The practical implementation of artificial intelligence technologies in medicine. Nat Med. 2019;25(30–36).

9. Topol EJ. High-performance medicine: the convergence of human and artificial intelligence. Nat Med. 2019;25:44–56.

10. Orlowski A, Snow S, Humphreys H, Smith W, Jones RS, Ashton R, et al. Bridging the impactibility gap in population health management: a systematic review. BMJ Open. 2021;11:52455.

11. Mora J, Iturralde MD, Prieto L, Domingo C, Gagnon MP, Martínez-Carazo C, et al. Key aspects related to implementation of risk stratification in health care systems-the ASSEHS study. BMC Health Serv Res. 2017;17(1):1–8.

12. JADECARE (2020-2023). Joint Action on implementation of digitally enabled integrated person-centred care [Internet]. 2020. Available from: https://www.jadecare.eu/

13. Deparment of Health. Government of Catalonia Health plan for 2011-2015. 2012.

14. 3M^TM^ Clinical Risk Groups (CRGs) | 3M [Internet]. [cited 2023 Apr 24]. Available from: https://www.3m.com/3M/en_US/health-information-systems-us/drive-value-based-care/patient-classification-methodologies/crgs/

15. Santaeugènia SJ, Contel JC, Vela E, Cleries M, Amil P, Melendo-Azuela EM, et al. Characteristics and Service Utilization by Complex Chronic and Advanced Chronic Patients in Catalonia: A Retrospective Seven-Year Cohort-Based Study of an Implemented Chronic Care Program. Int J Environ Res Public Heal 2021, Vol 18, Page 9473. 2021 Sep;18(18):9473.

16. Catalan Health Service. Catalan model of care for people with frailty, complex chronic (CCP) and advanced chronic (ACP) conditions. 2021.

17. Constante Beitia C. The Health Plan for Catalonia: an instrument to transform the health system. Med Clin (Barc). 2015 Nov 1;145 Suppl:20–6.

18. Monterde D, Vela E, Clèries M. Adjusted morbidity groups: A new multiple morbidity measurement of use in Primary Care. Aten primaria. 2016 Dec 1;48(10):674–82.

19. Monterde D, Vela E, Clèries M, García Eroles L, Pérez Sust P. Validity of adjusted morbidity groups with respect to clinical risk groups in the field of primary care. Aten Primaria. 2018 Feb 9;51(3):153–61.

20. Catalan Health System Observatory. Results Centre [Internet]. 2023 [cited 2023 Jun 8]. Available from: https://observatorisalut.gencat.cat/en/central_de_resultats/index.html

21. Catalan Health Department. Shared Clinical History in Catalonia [Internet]. [cited 2023 Jun 16]. Available from: https://salutweb.gencat.cat/ca/ambits_actuacio/linies_dactuacio/tic/sistemes-informacio/gestio-assistencial/hc3/index.html#googtrans(ca%7Cen)

22. Department of Health. Goverment of Catalonia Health plan for 2016–2020. 2016.

23. Ministerio de Sanidad Consumo y Bienestar Social. Informe del proyecto de Estratificación de la Población por Grupos de Morbilidad Ajustados (GMA) en el Sistema Nacional de Salud (2014-2016). Sistema Nacional de Salud. 2018.

24. Marin-Gomez FX, Mendioroz-Peña J, Mayer MA, Méndez-Boo L, Mora N, Hermosilla E, et al. Comparing the clinical characteristics and mortality of residential and non-residential older people with COVID-19: Retrospective observational study. Int J Environ Res Public Health. 2022 Jan 1;19(1):483.

25. Roso-Llorach A, Serra-Picamal X, Cos FX, Pallejà-Millán M, Mateu L, Rosell A, et al. Evolving mortality and clinical outcomes of hospitalized subjects during successive COVID-19 waves in Catalonia, Spain. Glob Epidemiol. 2022 Dec 1;4:100071.

26. Monterde D, Vela E, Clèries M, Garcia-Eroles L, Roca J, Pérez-Sust P. Multimorbidity as a predictor of health service utilization in primary care: a registry-based study of the Catalan population. BMC Fam Pract 2020 211. 2020 Feb 17;21(1):1–9.

27. Vela E, Piera-Jiménez J. Performance of Quantitative Measures of Multimorbidity: A Population-Based Retrospective Analysis. 2021 Mar 1;

28. Barrio-Cortes J, Castaño-Reguillo A, Beca-Martínez MT, Bandeira-de Oliveira M, López-Rodríguez C, Jaime-Sisó MÁ. Chronic diseases in the geriatric population: morbidity and use of primary care services according to risk level. BMC Geriatr. 2021 Dec 1;21(1):1–11.

29. Barrio-Cortes J, Soria-Ruiz-Ogarrio M, Martínez-Cuevas M, Castaño-Reguillo A, Bandeira-de Oliveira M, Beca-Martínez MT, et al. Use of primary and hospital care health services by chronic patients according to risk level by adjusted morbidity groups. BMC Health Serv Res. 2021 Dec 1;21(1):1–13.

30. Vela E, Tényi Á, Cano I, Monterde D, Cleries M, Garcia-Altes A, et al. Population-based analysis of patients with COPD in Catalonia: a cohort study with implications for clinical management. BMJ Open. 2018 Mar 1;8(3):e017283.

31. Department of Health. Generalitat de Catalunya. The Catalan Information Systems Master Plan Building a digital health strategy for Catalonia together. 2019.

32. Monterde D, Cainzos-Achirica M, Cossio-Gil Y, García-Eroles L, Pérez-Sust P, Arrufat M, et al. Performance of Comprehensive Risk Adjustment for the Prediction of In-Hospital Events Using Administrative Healthcare Data: The Queralt Indices. Risk Manag Healthc Policy. 2020;13:271.

33. Monterde D, Carot-Sans G, Cainzos-Achirica M, Abilleira S, Coca M, Vela E, et al. Performance of Three Measures of Comorbidity in Predicting Critical COVID-19: A Retrospective Analysis of 4607 Hospitalized Patients. 2021;

34. Goldfield N. The evolution of diagnosis-related groups (DRGs): From its beginnings in case-mix and resource use theory, to its implementation for payment and now for its current utilization for quality within and outside the hospital. Qual Manag Health Care. 2010 Jan;19(1):3–16.

35. ICD - ICD-9-CM - International Classification of Diseases, Ninth Revision, Clinical Modification [Internet]. [cited 2023 May 28]. Available from: https://www.cdc.gov/nchs/icd/icd9cm.htm

36. ICD - ICD-10-CM - International Classification of Diseases, Tenth Revision, Clinical Modification [Internet]. [cited 2021 Jul 21]. Available from: https://www.cdc.gov/nchs/icd/icd10cm.htm

37. Kindig D. What Are We Talking About When We Talk About Population Health? [Internet]. Health Affairs Forefront. 2015 [cited 2023 May 26]. Available from: https://www.healthaffairs.org/content/forefront/we-talking-we-talk-population-health

38. Christoff P. Running PDSA cycles. Curr Probl Pediatr Adolesc Health Care. 2018 Aug 1;48(8):198–201.

39. Baltaxe E, Hsieh HW, Roca J, Cano I. The Assessment of Medical Device Software Supporting Health Care Services for Chronic Patients in a Tertiary Hospital: Overarching Study. J Med Internet Res 2023;25e40976 https://www.jmir.org/2023/1/e40976.2023 Jan 4;25(1):e40976.

40. Herranz C, Martín L, Dana F, Sisó-Almirall A, Roca J, Cano I. Health Circuit: a practice-proven adaptive case management approach for innovative healthcare services. medRxiv. 2023 Mar 28;2023.03.22.23287569.

41. Bansback N, Bell M, Spooner L, Pompeo A, Han PKJ, Harrison M. Communicating Uncertainty in Benefits and Harms: A Review of Patient Decision Support Interventions. Patient. 2017 Jun 1;10(3):311–9.

42. Civita M, Roman R. PADRIS, una eina per a la millora dels processos assistencials, la recerca biomèdica i la planificació sanitària. Scientia. 2019;

43. Murray SA, Kendall M, Boyd K, Sheikh A. Illness trajectories and palliative care. BMJ. 2005 Apr 28;330(7498):1007–11.

44. Jensen AB, Moseley PL, Oprea TI, Ellesøe SG, Eriksson R, Schmock H, et al. Temporal disease trajectories condensed from population-wide registry data covering 6.2 million patients. Nat Commun. 2014 Jun 24;5(1):1–10.

