## Supplementary Material for "Toward adoption of health risk assessment in population-based and clinical scenarios"

Rubèn González-Colom et al.

*(On-line supplementary material)*

### 1 APPENDIX 1 RESULTS OF THE IMPLEMENTATION

#### 1.1 MARCHE REGION (I)

##### 1.1.1 Local Action Plan

###### *1.1.1.1 Local Good Practice*

<table>
<tr><td colspan="2">Local Good Practice</td><td colspan="2">A stratification tool for an effective management of chronic diseases in the Marche region</td></tr>
<tr><td colspan="3">Target population</td><td>Setting(s)</td></tr>
<tr><td colspan="3">The entire population of Marche region (~1,500,000 habitants)</td><td>The Regional Health System</td></tr>
<tr><td colspan="4">Main aim</td></tr>
<tr><td colspan="4">To apply a population stratification tool to improve the efficiency of the Regional Health System and the quality of life of citizens by providing services that meet their needs.</td></tr>
<tr><td>Outcomes</td><td colspan="2">Local Core Features and their Components</td><td>Inputs</td></tr>
<tr><td>• Providing an in-depth analysis of the health status of the Marche population<br>• Guaranteeing continuity of care throughout the region<br>• Reorganising services based on the health needs of people suffering from chronic diseases.<br>• Assessing the economic impact of the reorganisation of services for chronic diseases.</td><td colspan="2">Implement a risk stratification tool based on adjusted morbidity groups (LCF1)<br>• criteria for local stratification model<br>• data extraction and processing mechanisms<br>• definition of care programs for each of the strata<br>Build a map/dashboard of citizens' health/risk and available services (LCF2)<br>• City/village/district indicators based on the GMA and/or chronic diseases.<br>• Map available services in each area<br>• dashboard for data visualization</td><td>• Data<br>• Staff<br>• IT system<br>• Funding<br>• Decision-makers<br>• Technical assistance</td></tr>
<tr><td colspan="4">General description</td></tr>
<tr><td colspan="4">The intervention consists of setting up and testing a stratification tool for planning and decision-making purposes.</td></tr>
<tr><td colspan="4">Local Core Feature 1</td></tr>
<tr><td colspan="4">Component 1 is represented by the Catalan GMA population stratification algorithm, suitably adapted to the regional context and available health data. This tool will focus on chronic diseases and will make it possible to assess the population of the Marche Region placed in the higher sections of the Kaiser pyramid.</td></tr>
<tr><td colspan="4">Local Core Feature 2</td></tr>
<tr><td colspan="4">Component 2 envisages displaying on Marche region map the aggregated data from the stratification (and other indicators related to chronic diseases) as well as the available services to facilitate analysis and planning activities.</td></tr>
</table>

*1.1.1.2 Local Action Plan*

<table>
<tr><td>Local Good Practice</td><td colspan="5">A stratification tool for an effective management of chronic diseases in the Marche region</td></tr>
<tr><td colspan="4">Target population</td><td colspan="2">Setting</td></tr>
<tr><td colspan="4">The entire population of Marche region (~1,500,000 habitants)</td><td colspan="2">The Regional Health System</td></tr>
<tr><td colspan="6">Main aim</td></tr>
<tr><td colspan="6">To apply a population stratification tool to improve the efficiency of the Regional Health System and the quality of life of citizens by providing services that meet their needs.</td></tr>
<tr><td colspan="6">General description</td></tr>
<tr><td colspan="6">The intervention consists of setting up and testing a stratification tool for planning and decision-making purposes.</td></tr>
<tr><td>Related oGPs and CFs</td><td colspan="5">Catalan Open Innovation Hub on ICT<br>Block 1: Health Risk Assessment:<br>CF1.1 Assessment of transferability, and identification of steps for adoption, according to intellectual property rules, of the Catalan population-based risk stratification tool (AMG) into the ecosystem of the next adopter.<br>CF1.2 Health data management strategies<br>CF1.3 Development of enhanced risk prediction modelling for health policy purposes and/or clinical risk prediction.</td></tr>
<tr><td>Local Core Feature 1</td><td colspan="5">Implement a risk stratification tool based on adjusted morbidity groups (AMG)</td></tr>
<tr><td colspan="6">SMART objective</td></tr>
<tr><td colspan="6">In the framework of JADECARE, Marche Region will set up a risk stratification tool based on GMA, which will support healthcare services programming, with a focus on chronic diseases.</td></tr>
<tr><td>Activities</td><td>Actors</td><td>Resources</td><td>Setting(s)</td><td>Timeline</td><td>KPIs</td></tr>
<tr><td>Identify criteria for local stratification model</td><td>-Healthcare professionals<br>-IT experts<br>-Data scientists</td><td>Catalan GMA</td><td>Marche Region/Regional Health Agency</td><td>3 months (December 2021-February 2022)</td><td>List of criteria (Y/N)</td></tr>
<tr><td>Implement data extraction and processing mechanisms</td><td>-Healthcare professionals<br>-IT experts<br>-Data scientists</td><td>-Health data<br>-IT infrastructure<br>-Software</td><td>Marche Region/Regional Health Agency</td><td>6 months (March-August 2022)</td><td>-Database with required health data (Y/N)<br>-Availability of IT infrastructure (Y/N)<br>-% of data processed</td></tr>
<tr><td>Define care programs and services for each of the strata</td><td>-Healthcare professionals<br>-Marche/ARS staff</td><td>NA</td><td>Marche Region/Regional Health Agency</td><td>4 months (July-September 2022)</td><td>Short report on care programs and services</td></tr>
<tr><td>Local Core Feature 2</td><td colspan="5">Build a map/dashboard of citizens' health/risk and available services</td></tr>
<tr><td colspan="6">SMART objective</td></tr>
<tr><td colspan="6">In the framework of JADECARE, Marche Region will develop a map/dashboard allowing in-depth analysis of stratification data and available services for policy- and decision-making processes</td></tr>
<tr><td>Activities</td><td>Actors</td><td>Resources</td><td>Setting(s)</td><td>Timeline</td><td>KPIs</td></tr>
</table>

| | | | | | |
|---|---|---|---|---|---|
| Definition of indicators on stratification and/or chronic diseases | -Healthcare professionals<br>-IT experts<br>-Data scientists<br>-Marche/ARS staff | NA | Marche Region/Regional Health Agency | 2 months (September-October 2022) | List of indicators (Y/N) |
| Mapping of available services for chronic diseases in Marche Region | -Healthcare professionals<br>-Marche/ARS staff | Data on available services | Marche Region/Regional Health Agency | 2 months (September-October 2022) | List of services (Y/N) |
| Set up of the dashboard for data visualization | -Healthcare professionals<br>-IT experts<br>-Data scientists<br>-Marche/ARS staff | Subcontract for the dashboard | Marche Region/Regional Health Agency | 6 months (June-November 2022) | Availability of the dashboard (Y/N) |
| Identification of policies and interventions at regional level to support implementation and sustainability of the LGP | -Project manager<br>-Marche/ARS staff<br>-Regional policy representatives | NA | Marche Region/Regional Health Agency | 2022-2023 | List of policies and interventions (Y/N) |

##### 1.1.2 Plan-Do-Study-Act Cycles

###### *1.1.2.1 1st PDSA Cycle*

###### 1.1.2.1.1 Plan

| **LCF1** | **Implement a risk stratification tool based on adjusted morbidity groups (AMG)** | | | | | | | |
|---|---|---|---|---|---|---|---|---|
| **Activities** | **Actions** | **Actors** | **Timeline** | **KPIs measure (data collection)** | | | | |
| | | | | **KPI** | **Who** | **When** | **How** | **Target** |
| Activity 1: Identify criteria for local stratification model | Define the **cohort study** based on chronicity, comorbidities and vulnerability | -Computer specialist<br><br>-IT experts<br><br>-Healthcare professional<br><br>-Data scientists | 1/12/21 to 31/03/22 | List of criteria used for the stratification (Y/N) | Project Manager | 31/03/22 | Utilising information technology systems<br><br>Monitoring during the monthly follow-up meetings | Yes |
| | Define sources of stratification criteria:<br><br>1.ICD-9CM and ICD-10CM, from administrative health databases: Drugs, Hospital admission, Emergency and Urgency, Homecare, Exemption, Mortality registry.<br><br>2. National and regional criteria of extremely frail and vulnerable patients, in Covid-vaccination booking | | | | | | | |
| | Analysis of algorithm's preliminary results (strings' control and data validation) | | | | | | | |

| | | | | | | | | |
|---|---|---|---|---|---|---|---|---|
| Activity 2: Implement data extraction and processing mechanisms | Standardize variables format based on algorithm for stratification | -Computer specialist<br><br>-IT experts<br><br>-Healthcare professional<br><br>-Data scientists | 01/03/22 to 30/04/22 | -Database with required health data (Y/N)<br><br>-Availability of IT infrastructure (Y/N)<br><br>-% of data processed | Project manager | 30/04/22 | Utilising information technology systems<br><br>Monitoring during the monthly follow-up meetings | Yes |
| | Automating the data extraction and validation process | | 01/05/22 to 31/07/22 | | | 31/07/22 | | |
| | Implement algorithm tool | | 01/08/22 to 31/08/22 | | | 31/08/22 | | |
| Activity 3: Define care programs and services for each of the strata | Elaborate population management strategies for GMA 1,2,3,4, starting with health promotion, to case management strategy, focusing on chronic diseases management | -Healthcare professional<br><br>-Marche/ARS staff | 31/07/22 to 30/09/22 | Short report on care programs and services (Y/N) | Project manager | 30/09/22 | Relying on specific evaluation and guide from healthcare professionals<br><br>Monitoring during the monthly follow-up meetings | Yes |

#### 1.1.2.1.2 Do

| Cycle number (1 or 2) | 1 | |
|---|---|---|
| **Activity** | **KPI** | **Actual value** |
| **LCF 1 – Activity 1:** Identify criteria for local stratification model | List of criteria used for the stratification (Y/N) | Yes |
| **LCF 1 – Activity 2:** Implement data extraction and processing mechanisms | Database with required health data (Y/N) | No |
| | Availability of IT infrastructure (Y/N) | N.A.<br>(Expected during PDSA 2) |
| | % of data processed | N.A.<br>(Expected during PDSA 2) |
| **LCF 1 – Activity 3:** Define care programs and services for each of the strata | Short report on care programs and services (Y/N) | N.A.<br>(Expected during PDSA 2) |

| QUESTIONS | ANSWERS |
|---|---|
| **What was actually implemented? Any deviation from the planned actions** | Main achievements of PDSA1 are:<br>- identification of data sources as input for the GMA<br>- evaluation of the quality of the health data of Marche region<br>- evaluation of the applicability of the GMA in Marche Region<br>- preliminary analysis of the health status of Marche citizens.<br><br>Some deviations occurred: there were some delays due to the internal reorganisation and administrative procedures for personnel recruitment. Transfer of the algorithm and related documentation took more time than expected due to authorization procedures at the oGP site. The order of actions under Activity 2 changed after technical evaluations. |
| **Problems? Unexpected findings? Please describe** | Some problems/challenges occurred due to the internal reorganisation and the overburdened personnel. Access and management of the administrative health databases is challenging due to privacy restrictions, authorizations, IT issues. However, these issues are constantly monitored and partially solved.<br><br>Main barrier to the full implementation of the tool is the GDPR application in Italy.<br><br>As regards unexpected findings, we can acknowledge the great results obtained during the pilot test with the GMA. |

| IMPLEMENTATION PROGRESS OF THE LOCAL GOOD PRACTICE | | | |
|---|---|---|---|
| **0-25%** | **25-50%** | **50-75%** | **75-100%** |
| | **x** | | |

###### 1.1.2.1.3 Study

| **Cycle number (1or 2)** | | **1** | | | | |
|---|---|---|---|---|---|---|
| **Activity** | **KPI** | **Target value** | **Actual value** | **Reasons for the deviations** | **Mitigation actions implemented** | **Impact of mitigation actions** |
| **LCF 1 – Activity 1:** Identify criteria for local stratification model | List of criteria used for the stratification (Y/N) | Yes | Yes | | | |
| **LCF 1 – Activity 2:** Implement data extraction and processing mechanisms | Database with required health data (Y/N) | Yes | No | Some delay due to the internal reorganisation and administrative procedures for personnel recruitment. Transfer of the algorithm and related documentation took more time than expected due to authorization procedures at the oGP site. The order of actions under Activity 2 changed after technical evaluations | Monitoring of the recruitment procedures and collaboration with the administrative offices to speed up the process. Reorder of the activities while waiting the algorithm. | Personnel was successfully recruited. The new order of the activities increased the efficiency of the plan. |
| | Availability of IT infrastructure (Y/N) | Yes | N.A. (Expected during PDSA 2) | | | |
| | % of data processed | 100% | N.A. (Expected during PDSA 2) | | | |
| **LCF 1 – Activity 3:** Define care programs and services for each of the strata | Short report on care programs and services (Y/N) | Yes | N.A. (Expected during PDSA 2) | Not yet started | | |

###### 1.1.2.1.4 Act

| **Cycle number (1 or 2)** | 1 | | |
|---|---|---|---|
| **Activity** | **Maintain** | **Adapt** | **Abandon** |
| **LCF 1 – Activity 1:** Identify criteria for local stratification model | X | | |

| | | |
|---|---|---|
| **LCF 1 – Activity 2:** Implement data extraction and processing mechanisms | | X - The order of the actions needs to be changed due to technical |
| **LCF 1 – Activity 3:** Define care programs and services for each of the strata | | X - Timing of the actions needs to be extended. Alignment with National/regional guidelines currently in progress is needed. |

| QUESTIONS | ANSWERS |
|---|---|
| **Any new proposed action for the future?** | Pilot testing of GMA implementation |

##### *1.1.2.2 2nd PDSA Cycle*

###### 1.1.2.2.1 Plan

| **LCF1** | **Implement a risk stratification tool based on adjusted morbidity group (GMA)** | | | | | | | |
|---|---|---|---|---|---|---|---|---|
| **Activities (from the LAP)** | **Actions** | **Actors** | **Timeline** | **KPIs measure (data collection)** | | | | |
| | | | | **KPI** | **Who** | **When** | **How** | **Target** |
| Activity 1: Implement data extraction and processing mechanisms | Automating the data extraction and validation process | -Data analysts<br>-IT experts<br>-Technicians<br>Healthcare professionals | 01/04/22 to 31/08/22 | 1.Database with required health data (Y/N)<br>2.Short report on quality check (Y/N)<br>3.Procedure for data extraction (Y/N)<br>4. Short report on pilot test results (Y/N)<br>5.Integration of the tool in the regional IT infrastructure (Y/N) | Project manager<br>Data Analyst | 31/08/2022 | Utilising information technology systems<br>Monitoring during the weekly/monthly follow-up meetings | 1.Yes<br>2.Yes<br>3.Yes<br>4.Yes<br>5.Yes |
| | Standardize variables format based on algorithm for stratification | | | | | 30/09/2022 | | |
| | Pilot test of algorithm | | | | | 30/11/2022 | | |
| | Implement algorithm tool | | | | | 31/01/2023 | | |
| Activity 2: Define care programs and services for each of the strata | Elaborate population management strategies for GMA strata, starting with health promotion, to case management strategy, focusing on | -Healthcare professionals<br>-Marche/ARS staff | 01/09/22 to 30/11/22 | Guidelines on population management strategies (Y/N) | Project manager | 30/11/22 | Relying on specific evaluation and guidance from healthcare professionals Monitoring during the | Yes |

| | | | | | | | |
|---|---|---|---|---|---|---|---|
| | chronic diseases management | | | | | | monthly follow-up meetings |

| LCF2 | Build a map/dashboard of citizens' health/risk and available services | | | | | | | |
|---|---|---|---|---|---|---|---|---|
| **Activities (from the LAP)** | **Actions** | **Actors** | **Timeline** | **KPIs measure (data collection)** | | | | |
| | | | | **KPI** | **Who** | **When** | **How** | **Target** |
| Activity 1: Definition of indicators on stratification and/or chronic diseases | Mapping of relevant indicators at National/Regional levels | -Healthcare professionals<br>-Data analysts<br>-IT experts<br>-Marche/ARS staff | 01/09/22 to 30/09/22 | List of indicators (Y/N) | Project manager | 30/09/22<br>31/10/22 | Monitoring during the weekly/monthly follow-up meetings | Yes |
| | Development of new/revision of existing indicators | | 01/10/22 to 31/10/22 | | | | | |
| Activity 2: Mapping of available services for chronic diseases in Marche Region | Collection of databases/registries of health and social care facilities/services (e.g. community services, hospitals, residential and semi-residential facilities), including info on sources, timing of updates, classification and users | Marche/ARS staff | 01/09/22 to 30/11/22 | List of facilities/services (Y/N) | Project manager | 30/11/22 | Monitoring during the weekly/monthly follow-up meetings | Yes |
| Activity 3: Set up of the dashboard for | Revision of available platforms and definition of technical requirements | -Marche/ARS staff | 01/07/22 to 30/09/22 | Availability of the dashboard (Y/N) | Project manager | 31/12/22 | Monitoring during the | Yes |

| | | | | | | | | |
|---|---|---|---|---|---|---|---|---|
| data visualization | Implementation of the dashboard | -Healthcare professionals<br>-Data analysts<br>-IT experts | 01/10/22 to 31/12/22 | | | | weekly/monthly follow-up meetings | |
| Activity 4: Identification of policies and interventions at regional level to support implementation and sustainability of the LGP | Monitoring and analysis of relevant National/Regional guidelines, laws, plans | -Marche/ARS staff<br>-Healthcare professionals<br>-Data analysts<br>-Regional policy representatives | 01/04/22 to 30/09/23 | Sustainability plan (Y/N) | Project manager | 31/12/22 | Monitoring during the monthly follow-up meetings | Yes |
| | Definition of a sustainability plan for the integration of the tool in the decision-making process | | 01/09/22 to 31/12/22 | | | | | |

1.1.2.2.2 Do

| Cycle number (1 or 2) | 2 | |
|---|---|---|
| **Activity** | **KPI** | **Actual value** |
| LCF 1 - Activity 1: Implement data extraction and processing mechanisms | Database with required health data (Y/N) | Yes |
| | Short report on quality check (Y/N) | Yes |
| | Procedure for data extraction (Y/N) | Yes |
| | Short report on pilot test results (Y/N) | Yes |
| | Integration of the tool in the regional IT infrastructure (Y/N) | Yes (partially) |
| LCF 1 - Activity 2: Define care programs and services for each of the strata | Guidelines on population management strategies (Y/N) | Yes |
| LCF 2 - Activity 1: Definition of indicators on stratification and/or chronic diseases | List of indicators (Y/N) | Yes |
| LCF 2 - Activity 2: Mapping of available services for chronic diseases in Marche Region | List of facilities/services (Y/N) | Yes |
| LCF 2 - Activity 3: Set up of the dashboard for data visualization | Availability of the dashboard (Y/N) | No (Ongoing, preliminary structure of the dashboard available) |
| LCF 2 - Activity 4: Identification of policies and interventions at regional level to support implementation and sustainability of the LGP | Sustainability plan (Y/N) | Yes |

| QUESTIONS | ANSWERS |
|---|---|
| **What was actually implemented? Any deviation from the planned actions** | Main achievements of PDSA2 are:<br><br>- complete database of regional population, through the linkage of administrative health databases (Hospital discharges, |

| | |
|---|---|
| | Emergency & Urgency, Home care, Exemptions, population registry)<br>- quality evaluation of health data with identification of improvement actions (through quality reports and experts' opinion)<br>- report on population stratification, gained through GMA's first application.<br>- structure of a dashboard for indicators and data visualization<br>- guidelines of population management's strategies, through new National/Regional regulations<br><br>Some deviations occurred: there were some delays due to the complexity of the data preparation phase. Administrative health care databases' analysis and linkage took more time than expected due to their complexity and to the need of conducting data quality evaluation with health databases' experts. For this reason, the integration with the Regional IT system was delayed but partially achieved (completion expected in one month). Ongoing reorganisation of the regional healthcare system had an impact on some tasks due to overburdened personnel and for the ongoing changes in the IT systems.<br><br>The setting up of dashboard is complex, in terms of statistical computing and clinical relevance. Previous plans on reusing internal tools were abandoned (for the time being) because of changes in the organisation and time restrictions. Dashboard and indicators are being defined, also with the support of the oGP but the implementation is still ongoing, and it will continue during the post-implementation phase. |
| **Problems? Unexpected findings? Please describe** | Some challenges occurred due to the data complexity. Indeed, the management of the administrative health databases is challenging due to privacy restrictions, authorizations, IT issues. However, these issues are constantly monitored and partially solved. Main barrier to the full implementation of the tool is the GDPR application in Italy, still under debate.<br><br>As regards unexpected findings, we can acknowledge the great interest of healthcare managers in the project and their fruitful collaboration during the quality check and indicators/dashboard development phases. Indeed, quality data evaluation showed the need to conduct further analysis and insights of healthcare databases. Moreover, the fruitful discussions enhanced the identification of relevant aspects for healthcare services programming which will be considered in the next implementation period.<br><br>The data preparation phase was very challenging but, in the end, the results achieved confirmed the great potential of the tool. Moreover, we succeed to include the preliminary results of the algorithm in a Regional Deliberation on the adoption of the new |

| | |
|---|---|
| | National standards of community healthcare services, as a part of the regional implementation of the Recovery and Resilience Plan. |

| **IMPLEMENTATION PROGRESS OF THE LOCAL GOOD PRACTICE** | | | |
|---|---|---|---|
| **0-25%** | **25-50%** | **50-75%** | **75-100%** |
| | | | **x** |

###### 1.1.2.2.3 Study

| **Cycle number** | | **2** | | | | |
|---|---|---|---|---|---|---|
| **Activity** | **KPI** | **Target value** | **Actual value** | **Reasons for the deviations** | **Mitigation actions implemented** | **Impact of mitigation actions** |
| LCF 1 - Activity 1: Implement data extraction and processing mechanisms | Database with required health data (Y/N) | Yes | Yes | | | |
| | Short report on quality check (Y/N) | Yes | Yes | | | |
| | Procedure for data extraction (Y/N) | Yes | Yes | | | |
| | Short report on pilot test results (Y/N) | Yes | Yes | | | |
| | Integration of the tool in the regional IT infrastructure (Y/N) | Yes | Yes (partially) | Delay due to longer time needed to complete previous activities and overburdened personnel | Reorganization of the activities | Gain time to set up the server |
| LCF 1 - Activity 2: | Guidelines on population | Yes | Yes | | | |

| | | | | | | |
|---|---|---|---|---|---|---|
| Define care programs and services for each of the strata | management strategies (Y/N) | | | | | |
| LCF 2 - Activity 1: Definition of indicators on stratification and/or chronic diseases | List of indicators (Y/N) | Yes | Yes | | | |
| LCF 2 - Activity 2: Mapping of available services for chronic diseases in Marche Region | List of facilities/services (Y/N) | Yes | Yes | | | |
| LCF 2 - Activity 3: Set up of the dashboard for data visualization | Availability of the dashboard (Y/N) | Yes | No (Ongoing, preliminary structure of the dashboard available) | Delays in the data preparation phase; need to revise previously planned plans | New option under development | Enhancing implementation of the activity |
| LCF 2 - Activity 4: Identification of policies and interventions at regional | Sustainability plan (Y/N) | Yes | Yes | | | |

| |
|---|
| level to support implementation and sustainability of the LGP |

1.1.2.2.4 Act

| Cycle number (1 or 2) | 2 | | |
|---|---|---|---|
| **Activity** | **Maintain** | **Adapt** | **Abandon** |
| LCF 1 - Activity 1: Implement data extraction and processing mechanisms | | **X -** Additional activities are foreseen in the following months to: revise and simplify data preparation procedure; complete integration into IT system | |
| LCF 1 - Activity 2: Define care programs and services for each of the strata | **X -** Activity completed. Need to be monitored according to reorganization of the healthcare system and new regulations on healthcare services. | | |
| LCF 2 - Activity 1: Definition of indicators on stratification and/or chronic diseases | **X -** Activity completed. Need to be monitored to address new needs of healthcare managers and policymakers according to National requirements and reorganization of the Regional healthcare system. | | |
| LCF 2 - Activity 2: Mapping of available services for chronic diseases in Marche Region | **X -** Activity completed. Need to be monitored according to reorganization of the healthcare system and new regulations on healthcare services. | | |
| LCF 2 - Activity 3: Set up of the dashboard for data visualization | | **X -** Timing of the actions needs to be extended, to analyse in detail different options of data | |

| | | |
|---|---|---|
| | | visualization and find the most suitable solution for our context. It is foreseen the prosecution of the activity in the following months. |
| LCF 2 - Activity 4: Identification of policies and interventions at regional level to support implementation and sustainability of the LGP | **X** - Activity continuously progressing. | |

| **QUESTIONS** | **ANSWERS** |
|---|---|
| **Any new proposed action for the future?** | TECHNICAL/OPERATIVE ACTIONS:<br>• *Improvements in technical and graphic part of the dashboard*, also through development of maps to visualize healthcare services that will be activated based on recent social and health planning regulations (at national and regional level).<br>• *Revision of Indicators in alignment with the needs of experts in the various healthcare sectors,* e.g. through regular updating, and evaluation of their presentation modality at a higher level of detail (if possible, according to GDPR regulation).<br>• *Addition of further healthcare databases for AMG and indicators' computing (e.g.: hospice database)*, to make the data more accurate and to add information potentially relevant.<br>• *Make it a useful tool for regional and clinical managers* for close monitoring of population's health status and resources consumption, with possible comparison and benchmarking activities.<br>• Implementing of the dashboard in order to comply with the requirements of recent national and regional regulations regarding the *stratification mandate.*<br><br>SUSTAINABILITY ACTIONS:<br>• Enhancing and maximising AMG and dashboard's potential through *definition of a roadmap* to enhance its use as a tool for regional and clinical managers, e.g., for programming and implementing healthcare policies and services, process of budget and resources application, supporting investment in prevention and continuity of care (such as territorial operative centres, community hospitals and houses, home care, family nursing, palliative care, and telemedicine)<br>• Defining further analysis and actions to improve the quality and completeness of healthcare data. |

| | |
|---|---|
| | Disseminating data culture in institutions, raising awareness of the health care professionals on proper reporting of information |

#### 1.2 Estonia (EE)

##### 1.2.1 Local Action Plan

###### *1.2.1.1 Local Good Practice*

<table>
<tr><td colspan="2">Local Good Practice</td><td colspan="2">A funding model for person-centred and integrated services</td></tr>
<tr><td colspan="3">Target population</td><td>Setting(s)</td></tr>
<tr><td colspan="3">~50,000 habitants</td><td>Viljandi county</td></tr>
<tr><td colspan="4">Main aim</td></tr>
<tr><td colspan="4">Improve the results of the health and quality of life of the population and increase the efficiency of the healthcare system through better planning and use of resources.</td></tr>
<tr><td>Outcomes</td><td colspan="2">Local Core Features and their Components</td><td>Inputs</td></tr>
<tr><td><ul><li>Cooperation between Viljandi Hospital and other service providers is carried out.</li><li>IT tools supporting integrated care funding modelling and risk stratification.</li><li>Funding model has been proposed in integrated care provision in Viljandi county.</li><li>Assessment feasibility of nationwide implementation of oGP-s.</li></ul></td><td colspan="2">Develop a funding model for person-centred and integrated services:<ul><li>Risk stratification model</li><li>Case finding</li><li>Value-based contracting and payment framework</li><li>Analytical model to execute the contract.</li></ul></td><td><ul><li>Assessment of transferability of OptiMedis framework.</li><li>Assessment of transferability of risk stratification and case finding tools.</li><li>Identification of steps for adoption of the Catalan population-based risk stratification tool into the ecosystem of the NA.</li></ul></td></tr>
<tr><td colspan="4">General description</td></tr>
<tr><td colspan="4">Generating predictive model is needed to strengthen population health management and provide better-tailored services for risk groups. Contracting and funding models developed are lined with person-centred and integrated services.</td></tr>
<tr><td colspan="4">Local Core Feature 1</td></tr>
<tr><td colspan="4">Develop a funding model for person-centred and integrated services:<ul><li>Risk stratification model.</li><li>Case finding.</li><li>Value-based contracting and payment framework.</li><li>Analytical model to execute the contract.</li></ul></td></tr>
</table>

##### *1.2.1.2 Local Action Plan*

| **Local Good Practice** | A funding model for person-centred and integrated services |
|---|---|

| **Target population** | **Setting** |
|---|---|
| ~50,000 habitants | Viljandi county |

| **Main aim** |
|---|
| Improve the results of the health and quality of life of the population and increase the efficiency of the healthcare system through better planning and use of resources. |

| **General description** |
|---|
| Generating predictive model is needed to strengthen population health management and provide better-tailored services for risk groups. Contracting and funding models developed are lined with person-centred and integrated services. |

| | |
|---|---|
| **Related oGPs and CFs** | Mix'n'Match OptiMedis & Catalan OGP<br><br>CF1.1 Assessment of transferability, and identification of steps for adoption, according to intellectual property rules, of the Catalan population-based risk stratification tool (AMG) into the ecosystem of the next adopter.<br>CF1.2 Health data management strategies<br>CF1.3 Development of enhanced risk prediction modelling for health policy purposes and/or clinical risk prediction |

| **Local Core Feature 1** | **Develop a funding model for person-centred and integrated services.** |
|---|---|

| **SMART objective** |
|---|
| We will design a contracting and payment framework approach based on OptiMedis that includes Catalonian AMG risk stratification model. |

| **Activities** | **Actors** | **Resources** | **Setting(s)** | **Timeline** | **KPIs** |
|---|---|---|---|---|---|
| Create a core group to define the local contracting and payment framework model | -GPs, nurses<br>-Hospital doctors and nurses<br>-Healthcare planning experts | Professionals from different settings | -GP practices<br>-Viljandi Hospital<br>-Estonian Ministry of Social Affairs | 01.01.2022,<br>(5 months) | Number and profile of professionals engaged in the definition of the contracting and payment framework approach. |
| Establish criteria for contracting | -Healthcare professionals | Professionals from | -Viljandi Hospital | 02.02.2022<br>(4 months) | List of criteria used for contracting |

| | | | | | |
|---|---|---|---|---|---|
| and payment framework | -OptiMedis experts<br>-AMG experts | different settings | -Estonian Ministry of Social Affairs<br>-EHIF | | and payment framework (Y/N) |
| Set up the data extraction and processing mechanisms | -IT experts<br>-Data scientists<br>-OptiMedis experts<br>-AMG experts | -OptiMedis experts<br>-AMG experts<br>-IT infrastructure<br>-Subcontractor for technical development | Viljandi Hospital | 01.01.2022<br>(5 months) | -Database creation (Y/N)<br>-Technical design (%)<br>-Functional design (%) |
| Implement case finding and risk stratification | -IT experts<br>-Data scientists<br>-OptiMedis experts<br>-AMG experts | -IT infrastructure<br>-Subcontractor for technical development | Viljandi Hospital | 01.04.2022<br>(5 months) | Case finding and risk stratification tool is implemneted (Y/N) |
| Design contracting and payment framework | -GPs<br>-Hospital management.<br>-OptiMedis experts<br>-AMG experts | Professionals from different settings | -Viljandi Hospital<br>-Estonian Ministry of Social Affairs<br>-EHIF | 01.04.2022<br>(5 months) | Contracting and payment framework agreed (Y/N) |

| | | | | | |
|---|---|---|---|---|---|
| Assess case finding and risk stratification ba sed contracting and payment framework agai nst established criteria | -Experts<br>-Healthcare professionals<br>-Healthcare planning experts | Profession als from different settings | - Viljandi Hos pital<br>-GP practices<br>-Estonian Ministry of Social Affairs<br>-EHIF | 01.09.2022<br>(3 months) | Conformance report (Y/N) |

##### 1.2.2 Plan-Do-Study-Act Cycles

###### *1.2.2.1 1st PDSA Cycle*

###### 1.2.2.1.1 Plan

| **LCF1** | **Develop a funding model for person-centred and integrated services.** | | | | | | | |
|---|---|---|---|---|---|---|---|---|
| **Activities (from the LAP)** | **Actions** | **Actors** | **Timeline** | **KPIs measure (data collection)** | | | | |
| | | | | **KPI** | **Who** | **When** | **How** | **Target** |
| Activity 1: Create a core group to define the local contracting and payment framework model | Reaching core stakeholders agreement | -GPs, nurses.<br><br>-Hospital doctors and nurses.<br><br>-Healthcare planning experts. | 01.01.22 - 31.01.22 | Number and profile of professionals engage d in the definition of the contracting and payment framework approach | VH project manager | On stakeholder' s meetings | Registration forms | 5 |
| | Expanding and agreeing with key stakeholders | -GPs, nurses.<br><br>-Hospital doctors and nurses.<br><br>-Healthcare planning experts | 01.02.22 - 28.02.22 | | | | | |
| | Agreeing on all stakeholder's letter of intent of Viljandi county | -GPs, nurses.<br><br>-Hospital doctors and nurses. | 01.03.22 - 31.03.22 | | | | | |

| | | | | | | | | |
|---|---|---|---|---|---|---|---|---|
| | | -Healthcare planning experts | | | | | | |
| Activity 2: Establish criteria for contracting and payment framework | Introduction and creating possible scenarios | -IT Experts<br><br>-Healthcare professionals | 02.02.22<br>-<br>28.02.22 | List of criteria used for contracting and payment framework (Y/N) | VH project manager | On workshop | Agreement | Yes |
| | Agreeing roadmap | -IT Experts<br><br>-Healthcare professionals | 01.03.22<br>-<br>31.03.22 | | | | | |
| | Defining alternatives | -IT Experts<br><br>-Healthcare professionals | 01.04.22<br>-<br>30.04.22 | | | | | |
| | Concluding agreement | -IT Experts<br><br>-Healthcare professionals | 01.05.22<br>-<br>31.05.22 | | | | | |
| Activity 3: Set up the data extraction and processing mechanisms. | Improving incrementally data extract and loading (ETL) according to model requirements - Phase 1 | -IT experts<br><br>-Data scientists | 01.01.22<br>-<br>31.01.22 | 1.Database creation (Y/N)<br><br>2.Technical design (%)<br><br>3.Functional design (%) | VH project manager | After Phase 5 data extract and loading | Phase 5 report on data extract and loading | 1.Yes<br><br>2.100%<br><br>3.100% |
| | Improving incrementally data extract and loading (ETL) - Phase 2 | -IT experts<br><br>-Data scientists | 01.02.22<br>-<br>28.02.22 | | | | | |

| | | | | | | | | |
|---|---|---|---|---|---|---|---|---|
| | Improving incrementally data extract and loading (ETL) - Phase 3 | -IT experts<br>-Data scientists | 01.03.22<br>-<br>31.03.22 | | | | | |
| | Improving incrementally data extract and loading (ETL) - Phase 4 | -IT experts<br>-Data scientists | 01.04.22<br>-<br>30.04.22 | | | | | |
| | Improving incrementally data extract and loading (ETL) - phase 5 | -IT experts<br>-Data scientists | 01.05.22<br>-<br>31.05.22 | | | | | |
| Activity 4: Implement case finding and risk stratification. | Gathering and systematising LAP specific information and planning further action based upon agreed roadmap | -IT experts<br>-Data scientists | 01.4.22-<br>30.4.22 | Case finding and risk stratification tool is implemented (Y/N) | VH project manager | After publishing local case finding and risk stratification framework | Published local case finding and risk stratification framework | Yes |
| | Drafting local case finding and risk stratification framework | -IT experts<br>-Data scientists | 01.05.22<br>-31.5.22 | | | | | |
| | Drafting local case finding and risk stratification framework | -IT experts<br>-Data scientists | 01.06.22<br>-<br>30.06.22 | | | | | |
| | Agreeing on local case finding and risk stratification framework | -IT experts<br>-Data scientists | 01.08.22<br>-<br>31.08.22 | | | | | |

| | | | | | | | | |
|---|---|---|---|---|---|---|---|---|
| Activity 5:<br>Design contracting and payment framework. | Gathering and systematising LAP specific information and planning further action based upon agreed roadmap | -IT Experts<br><br>-Healthcare professionals<br><br>-Healthcare planning experts | 01.4.22-30.4.22 | Contracting and payment framework proposed (Y/N) | VH project manager | After publishing local contracting and payment framework | Published local contracting and payment framework | Yes |
| | Drafting local contracting and payment framework | -IT Experts<br><br>-Healthcare professionals<br><br>-Healthcare planning experts | 01.05.22 -31.5.22 | | | | | |
| | Improving local contracting and payment framework | -IT Experts<br><br>-Healthcare professionals<br><br>-Healthcare planning experts | 01.06.22 - 30.06.22 | | | | | |
| | Agreeing on local contracting and payment framework | -IT Experts<br><br>-Healthcare professionals<br><br>-Healthcare planning experts | 01.08.22 - 31.08.22 | | | | | |

| | | | | | | | | |
|---|---|---|---|---|---|---|---|---|
| Activity 6:<br>Assess proposed case finding and risk stratification based contr acting and payment framework against estab lished criteria. | Introducing preliminary report and gathering feedback | -IT Experts<br><br>-Healthcare professionals<br><br>-Healthcare planning experts | 01.09.22<br>-<br>30.09.22 | Conformance report (Y/N) | VH project manager | After publishing stakeholder s contracting and payment framework model agreement | Published stakeholder s contracting and payment framework model agreement | Yes |
| | Introducing improved report and gathering feedback | -IT Experts<br><br>-Healthcare professionals<br><br>-Healthcare planning experts | 01.10.22<br>-<br>31.10.22 | | | | | |
| | Introducing agreement draft and gathering feedback and finalizing report and agreement | -IT Experts<br><br>-Healthcare professionals<br><br>-Healthcare planning experts | 01.11.22<br>-<br>30.11.22 | | | | | |

### 1.2.2.1.2 Do

| Cycle number (1 or 2) | 1 | |
|---|---|---|
| **Activity** | **KPI** | **Actual value** |
| **LCF 1 - Activity 1:** Create a core group to define the local contracting and payment framework model | Number and profile of professionals engaged in the definition of the contracting and payment framework approach | 5 |
| **LCF 1 - Activity 2:** Establish criteria for contracting and payment framework | List of criteria used for contracting and payment framework (Y/N) | Yes |
| **LCF 1 - Activity 3:** Set up the data extraction and processing mechanisms. | Database creation (Y/N) | Yes |
| | Technical design (%) | 100% |
| | Functional design (%) | 100% |
| **LCF 1 - Activity 4:** Implement case finding and risk stratification. | Case finding and risk stratification tool is implemented (Y/N) | To be completed in PDSA 2 |
| **LCF 1 - Activity 5**: Design contracting and payment framework. | Contracting and payment framework proposed (Y/N) | To be completed in PDSA 2 |
| **LCF 1 – Activity 6:** Assess proposed case finding and risk stratification based contracting and payment framework against established criteria. | Conformance report (Y/N) | To be completed in PDSA 2 |

| QUESTIONS | ANSWERS |
|---|---|
| **What was actually implemented? Any deviation from the planned actions** | Health care professionals from primary care & hospital care are engaged (GPs, specialist doctors, RN both family nurses and hospital and home care nurses, social workers, data manger), planned no (5) was exceeded for synergy of combined expertise. There is a list of criteria of the contracting and payment framework approach created. Database is created and design requirements for technical and functionality full-filled. |
| **Problems? Unexpected findings? Please describe** | Due to current situation in health care has slowed down the planned activities, meetings have been held, plans expanded and agreed with key stakeholders discussed. |

| IMPLEMENTATION PROGRESS OF THE LOCAL GOOD PRACTICE | | | |
|---|---|---|---|
| **0-25%** | **25-50%** | **50-75%** | **75-100%** |
| | **X** | | |

##### 1.2.2.1.3 Study

| Cycle number | | 1 | | | | |
|---|---|---|---|---|---|---|
| **Activity** | **KPI** | **Target value** | **Actual value** | **Reasons for the deviations** | **Mitigation actions implemented** | **Impact of mitigation actions** |

| | | | |
|---|---|---|---|
| **LCF 1 - Activity 1:** Create a core group to define the local contracting an d payment framework model | Number and profile of professionals en gaged in the definition of the contracting and payment framework appr oach | 5 | 5 |
| **LCF 1 - Activity 2:** Establish criteria for contracting and payment framework | List of criteria used for contracting and payment framework (Y/N ) | Yes | Yes |
| **LCF 1 - Activity 3:** Set up the data extraction and processing mechanisms. | Database creation (Y/N) | Yes | Yes |
| | Technical design (%) | 100% | 100% |
| | Functional design (%) | 100% | 100% |
| **LCF 1 - Activity 4:** Implement case finding and risk stratification. | Case finding and risk stratification tool is implemented (Y/N) | Yes | To be completed in PDSA 2 |
| **LCF 1 - Activity 5:** Design contracting and payment framework. | Contracting and payment framework proposed (Y/N) | Yes | To be completed in PDSA 2 |
| **LCF 1 – Activity 6:** Assess proposed case finding and risk stratification based contracting and payment framework against established criteria. | Conformance report (Y/N) | Yes | To be completed in PDSA 2 |

###### 1.2.2.1.4 Act

| **Cycle number (1 or 2)** | **1** | | |
|---|---|---|---|
| **Activity** | **Maintain** | **Adapt** | **Abandon** |

| | |
|---|---|
| **LCF 1 - Activity 1:** Create a core group to define the local contracting and payment framework model | **X** |
| **LCF 1 - Activity 2:** Establish criteria for contracting and payment framework | **X** |
| **LCF 1 - Activity 3:** Set up the data extraction and processing mechanisms. | **X** |
| **LCF 1 - Activity 4:** Implement case finding and risk stratification. | **X** |
| **LCF 1 - Activity 5:** Design contracting and payment framework. | **X** |
| **LCF 1 – Activity 6:** Assess proposed case finding and risk stratification based contracting and payment framework against established criteria. | **X** |

| QUESTIONS | ANSWERS |
|---|---|
| **Any new proposed action for the future?** | oGP expertise is present and available; national stakeholders are interested; discussions are in place to define synergies between partners and opportunities |

##### *1.2.2.2 2nd PDSA Cycle*

###### 1.2.2.2.1 Plan

| **LCF1** | **Develop a funding model for person-centred and integrated services.** | | | | | | | |
|---|---|---|---|---|---|---|---|---|
| **Activities (from the LAP)** | **Actions** | **Actors** | **Timeline** | **KPIs measure (data collection)** | | | | |
| | | | | **KPI** | **Who** | **When** | **How** | **Target** |
| Activity 1: Create a core group to define the local contracting and payment framework model | Reaching core stakeholders agreement | -GPs, nurses.<br><br>-Hospital doctors and nurses.<br><br>-Healthcare planning experts. | 01.01.22 - 31.01.22 | Number and profile of professionals engage d in the definition of the contracting and payment framework approach | VH project manager | On stakeholder's meetings | Registration forms | 5 |
| | Expanding and agreeing with key stakeholders | -GPs, nurses.<br><br>-Hospital doctors and nurses.<br><br>-Healthcare planning experts | 01.02.22 - 28.02.22 | | | | | |
| | Agreeing on all stakeholder's letter of intent of Viljandi county | -GPs, nurses.<br><br>-Hospital doctors and nurses. | 01.03.22 - 31.03.22 | | | | | |

| | | | | | | | | |
|---|---|---|---|---|---|---|---|---|
| | | -Healthcare planning experts | | | | | | |
| Activity 2: Establish criteria for contracting and payment framework | Introduction and creating possible scenarios | -IT Experts<br>-Healthcare professionals | 02.02.22 - 28.02.22 | List of criteria used for contracting and payment framework (Y/N) | VH project manager | On workshop | Agreement | Yes |
| | Agreeing roadmap | -IT Experts<br>-Healthcare professionals | 01.03.22 - 31.03.22 | | | | | |
| | Defining alternatives | -IT Experts<br>-Healthcare professionals | 01.04.22 - 30.04.22 | | | | | |
| | Concluding agreement | -IT Experts<br>-Healthcare professionals | 01.05.22 - 31.05.22 | | | | | |
| Activity 3: Set up the data extraction and processing mechanisms. | Improving incrementally data extract and loading (ETL) according to model requirements - Phase 1 | -IT experts<br>-Data scientists | 01.01.22 - 31.01.22 | 1.Database creation (Y/N)<br>2.Technical design (%)<br>3.Functional design (%) | VH project manager | After Phase 5 data extract and loading | Phase 5 report on data extract and loading | 1.Yes<br>2.100%<br>3.100% |
| | Improving incrementally data extract and loading (ETL) - Phase 2 | -IT experts<br>-Data scientists | 01.02.22 - 28.02.22 | | | | | |
| | Improving incrementally data extract and loading (ETL) - Phase 3 | -IT experts | 01.03.22 - 31.03.22 | | | | | |

| | | | | | | | | |
|---|---|---|---|---|---|---|---|---|
| | | -Data scientists | | | | | | |
| | Improving incrementally data extract and loading (ETL) - Phase 4 | -IT experts<br><br>-Data scientists | 01.04.22<br>-<br>30.04.22 | | | | | |
| | Improving incrementally data extract and loading (ETL) - phase 5 | -IT experts<br><br>-Data scientists | 01.05.22<br>-<br>31.05.22 | | | | | |
| Activity 4: Implement case finding and risk stratification. | Gathering and systematising LAP specific information and planning further action based upon agreed roadmap | -IT experts<br><br>-Data scientists | 01.4.22-<br>30.4.22 | Case finding and risk stratification tool is implemented (Y/N) | VH project manager | After publishing local case finding and risk stratificatio n framework | Published local case finding and risk stratificatio n framework | Yes |
| | Drafting local case finding and risk stratification framework | -IT experts<br><br>-Data scientists | 01.05.22<br>-31.5.22 | | | | | |
| | Drafting local case finding and risk stratification framework | -IT experts<br><br>-Data scientists | 01.06.22<br>-<br>30.06.22 | | | | | |
| | Agreeing on local case finding and risk stratification framework | -IT experts<br><br>-Data scientists | 01.08.22<br>-<br>31.08.22 | | | | | |
| | Gathering and systematising LAP specific information and | -IT Experts | 01.4.22-<br>30.4.22 | | | After publishing | Published local | Yes |

| | | | | | | | |
|---|---|---|---|---|---|---|---|
| Activity 5:<br>Design contracting and payment framework. | planning further action based upon agreed roadmap | -Healthcare professionals<br><br>-Healthcare planning experts | | Contracting and payment framework proposed (Y/N) | VH project manager | local contracting and payment framework | contracting and payment framework |
| | Drafting local contracting and payment framework | -IT Experts<br><br>-Healthcare professionals<br><br>-Healthcare planning experts | 01.05.22 -31.5.22 | | | | |
| | Improving local contracting and payment framework | -IT Experts<br><br>-Healthcare professionals<br><br>-Healthcare planning experts | 01.06.22 - 30.06.22 | | | | |
| | Improving local contracting and payment framework based on CPTS contracting and payment framework | -IT Experts<br><br>-Healthcare professionals<br><br>-Healthcare planning experts | 01.08.22 - 31.08.22 | | | | |

| | | | | | | | | |
|---|---|---|---|---|---|---|---|---|
| Activity 6:<br>Assess proposed case finding and risk stratification based contr acting and payment framework against estab lished criteria. | Introducing preliminary report and gathering feedback | -IT Experts<br><br>-Healthcare professionals<br><br>-Healthcare planning experts | 01.09.22<br>-<br>30.09.22 | Conformance report (Y/N) | VH project manager | After publishing stakeholder s contracting and payment framework model agreement | Published stakeholder s contracting and payment framework model agreement | Yes |
| | Introducing improved report and gathering feedback | -IT Experts<br><br>-Healthcare professionals<br><br>-Healthcare planning experts | 01.10.22<br>-<br>31.10.22 | | | | | |
| | Introducing agreement draft and gathering feedback.<br>Finalizing report and agreement | -IT Experts<br><br>-Healthcare professionals<br><br>-Healthcare planning experts | 01.11.22<br>-<br>30.11.22 | | | | | |

1.2.2.2.2 Do

| Cycle number (1 or 2) | **2** | |
|---|---|---|
| **Activity** | **KPI** | **Actual value** |
| **LCF 1 - Activity 1**: Create a core group to define the local contracting and payment framework model | Number and profile of professionals engaged in the definition of the contracting and payment framework approach | 5 |
| **LCF 1 - Activity 2:** Establish criteria for contracting and payment framework | List of criteria used for contracting and payment framework (Y/N) | Yes |
| **LCF 1 - Activity 3:** Set up the data extraction and processing mechanisms. | Database creation (Y/N) | Yes |
| | Technical design (%) | 100% |
| | Functional design (%) | 100% |
| **LCF 1 - Activity 4:** Implement case finding and risk stratification. | Case finding and risk stratification tool is implemented (Y/N) | Yes |
| **LCF 1 - Activity 5:** Design contracting and payment framework. | Contracting and payment framework proposed (Y/N) | Yes |
| **LCF 1 – Activity 6:** Assess proposed case finding and risk stratification based contracting and payment framework against established criteria. | Conformance report (Y/N) | Yes |

| QUESTIONS | ANSWERS |
|---|---|
| **What was actually implemented? Any deviation from the planned actions** | Case finding and risk stratification tool is used locally, sustainability actions are planned to implement the tool at national level (national project “PAIK2022-2025” initiated). Contracting and payment framework agreed among the current project team, discussions with stakeholders done and framework implementation will follow over some time of the period. |
| **Problems? Unexpected findings? Please describe** | Regarding the contracting and payment framework model implementation on municipality level - the local level stakeholders & collaborative partner active involvement is slightly slowed down due to the raised workload related to their usual tasks, and the change of contact person; stakeholders contracting and payment framework model agreement reporting delayed. |

| IMPLEMENTATION PROGRESS OF THE LOCAL GOOD PRACTICE |
|---|

| 0-25% | 25-50% | 50-75% | 75-100% |
|---|---|---|---|
| | | | **X** |

1.2.2.2.3 Study

| **Cycle number** | | **2** | | | | |
|---|---|---|---|---|---|---|
| **Activity** | **KPI** | **Target value** | **Actual value** | **Reasons for the deviations** | **Mitigation actions implemented** | **Impact of mitigation actions** |
| **LCF 1 - Activity 1:** Create a core group to define the local contracting and payment framework model | Number and profile of professionals engaged in the definition of the contracting and payment framework approach | 5 | 5 | | | |
| **LCF 1 - Activity 2:** Establish criteria for contracting and payment framework | List of criteria used for contracting and payment framework (Y/N) | Yes | Yes | | | |
| **LCF 1 - Activity 3:** Set up the data extraction and processing mechanisms. | Database creation (Y/N) | Yes | Yes | | | |
| | Technical design (%) | 100% | 100% | | | |
| | Functional design (%) | 100% | 100% | | | |
| **LCF 1 - Activity 4:** Implement case finding and risk stratification. | Case finding and risk stratification tool is implemented (Y/N) | Yes | Yes | | | |
| **LCF 1 - Activity 5:** Design contracting and payment framework. | Contracting and payment framework proposed (Y/N) | Yes | Yes | | | |
| **LCF 1 – Activity 6:** | Conformance report (Y/N) | Yes | Yes | | | |

| |
|---|
| Assess proposed case finding and risk stratification based contracting and payment framework against established criteria. |

1.2.2.2.4 Act

| Cycle number (1 or 2) | 2 | | |
|---|---|---|---|
| **Activity** | **Maintain** | **Adapt** | **Abandon** |
| **LCF 1 - Activity 1:** Create a core group to define the local contracting and payment framework model | **X** | | |
| **LCF 1 - Activity 2:** Establish criteria for contracting and payment framework | **X** | | |
| **LCF 1 - Activity 3:** Set up the data extraction and processing mechanisms. | **X** | | |
| **LCF 1 - Activity 4:** Implement case finding and risk stratification. | **X** | | |
| **LCF 1 - Activity 5:** Design contracting and payment framework. | **X** | | |
| **LCF 1 – Activity 6:** Assess proposed case finding and risk stratification based contracting and payment framework against established criteria. | **X** | | |

| QUESTIONS | ANSWERS |
|---|---|
| **Any new proposed action for the future?** | No completely new activities proposed but some oGP support while ensuring the sustainability (both on regional and national level) of already planned and tested activities might be beneficial. |

### APPENDIX 2: JADECARE THEMATIC WORKSHOP, LECTURE ON HEALTH RISK ASSESSMENT, DAVID MONTERDE - VILJANDI 14TH JUNE 2022

Link to access the recording of the lecture on health risk assessment given by David Monterde in Viljandi (14th June 2022) in the context of the health risk assessment workshop as part of the activity of the EU project JADECARE:

https://drive.google.com/drive/folders/1Dwx2C2Ek2Z3mSz-2MRCJepWmzDrEggdh?usp=sharing
